# Differences in body temperature drive sex-specific immune responses

**DOI:** 10.1101/2025.09.29.25336741

**Authors:** Elizabeth Maloney, Rémi Trimbour, Marwan Sharawy, Jamie Sugrue, Patrick Terrence Brooks, Etienne Villain, COSIPOP study group, Rebecka Svanberg Teglgaard, Maria Elizabeth Engel Møller, Birgitte Lindegaard, Anne Ortved Gang, Susanne Dam Nielsen, Daria Podlekareva, Carsten Utoft Niemann, Sisse Rye Ostrowski, COVIMUN study consortium, Maxime Rotival, Etienne Patin, Lluis Quintana-Murci, Darragh Duffy, Milieu Intérieur Consortium

## Abstract

Human body temperature is sexually dimorphic, with females averaging 0.1-0.5°C warmer than males. Body temperature is also linked to immune responses through highly conserved fever and heat shock pathways. We hypothesized that subtle temperature changes could mediate sex differences in immune responses, with potential clinical implications. To test this, we analyzed 975 healthy adults from the *Milieu Interieur* cohort, confirming sex and age effects on homeostatic body temperature. Within the narrow 36-38°C range, we observed sex-specific associations with baseline and induced immune responses. In males, higher temperature was associated with decreased interferon alpha responses following bacterial and viral stimulation, whereas in females, it correlated with decreased interferon gamma responses after superantigen stimulation. These findings were recapitulated at fever temperatures *ex vivo*, and validated clinically in COVID-19 patients, with male-specific febrile suppression of antiviral pathways. Multicellular network analysis of stimulated single-cell data identified cellular regulatory cascades coordinating these temperature-dependent effects. To our knowledge, this is the first systems-level study of how natural variation in body temperature contributes to immune variation underlying sex disparities in disease.

## Introduction

Sex is one of the strongest determinants of susceptibility to autoimmune, malignant, and infectious disease. Many autoimmune diseases are four times more likely in females, while non-reproductive cancers are twice as common in males^1^. Males are also generally more susceptible to bacterial and viral infections, while females mount stronger antibody responses to vaccination but experience more adverse effects^2,3^. Although societal gender roles contribute to these disparities^1,2^, population-immunology approaches have revealed the substantial effect of biological sex on immune responses^4^, with the underlying mechanisms still being elucidated. Contributing factors include X-linked inactivation escape^5,6^ and steroid hormone influences^7–9^; however, a complete understanding is lacking.

Body temperature is a sexually dimorphic factor with a known impact on immunity^10,11^. Temperature and immune responses are linked through highly conserved pathways such as heat shock response (HSR) and fever. Fever triggers HSR in both bacterial and human immune cells ^12–14^, as hyperthermia activates this stress response to maintain protein folding under oxidative stress. Heat shock proteins (HSPs) induced during this process can exert proinflammatory effects by engaging toll-like receptors (TLRs), or anti-inflammatory effects via interleukin (IL)-10 induction in regulatory T cells^15–19^. Accordingly, fever and hyperthermia can be either stimulatory or suppressive, depending on cohort demographics, disease context, and immune cell type^20–25^. However, specialized studies have failed to provide a unified systems-level understanding of how temperature modulates immunity. This gap is compounded by natural fluctuations in healthy body temperature, which varies between and within individuals across daily, menstrual, seasonal, and age-related cycles^10^.

We hypothesized that subtle fluctuations in body temperature affect immune responses and may contribute to sex-specific differences in immunity. To address these questions, we analyzed cellular, transcriptomic, and proteomic responses in 949 healthy donors from the Milieu Interieur (MI) cohort. We found sex-and pathogen-specific effects of homeostatic temperature variation on immune responses that mirror sex disparities in disease. We then performed *ex vivo* whole blood stimulations at both homeostatic and febrile temperatures in 26 healthy donors, revealing attenuated interferon gamma (IFN-γ) signaling at febrile temperatures. Single-cell transcriptomic analyses confirmed these findings and identified the most affected cell types. Finally, temperature attenuation of male viral immune responses was confirmed in 82 COVID-19 patients, and through multicellular network analysis we identified potential cellular regulatory cascades coordinating this modulation. Collectively, these studies provide new insights into how variation in body temperature may contribute to sex-specific immune function.

## Results

### Interindividual body temperature variation explained by age, sex, and lifestyle

To assess interindividual variability in body temperature within the MI cohort, we fit linear models to ear temperatures measured at the immunological sampling visit, and assessed associations between sex, age, and body temperature using a generalized linear model with a Gamma distribution (glmGamma) (**Figure 1A**). Female donors exhibited a higher average body temperature than males (β = −0.46), and body temperature decreased with age in both sexes (β = −0.0082) (**Figure 1A**). The age decline was more pronounced in females, as evidenced by an age×sex interaction (β = 0.0051), but this sex difference was eliminated when restricting analysis to menopausal females and age-matched males (n=192, β=0.0042). For the decade with both pre-menopausal and menopausal donors, menopause was associated with cooler body temperature (44 – 55 years, n=63, β=-0.0050) (**Figure 1B**). A subset of 500 donors had 3 consecutive temperature measurements over a span of 2 - 8 weeks (median 4 weeks) (**Figure 1C**). In this subcohort, the same results were observed after modeling the mean by age (β=-0.00026), sex (β= −0.015), and their interaction (β=0.00018). Dispersion formula results revealed female donors showed overall more variability in body temperature than males (β=0.25), likely due to fluctuations associated with the menstrual cycle^10^. This was further supported by pre-menopausal female donors having more variability in body temperature than their post-menopausal counterparts (β=0.61) (**Figure 1D**).

**Figure 1.**
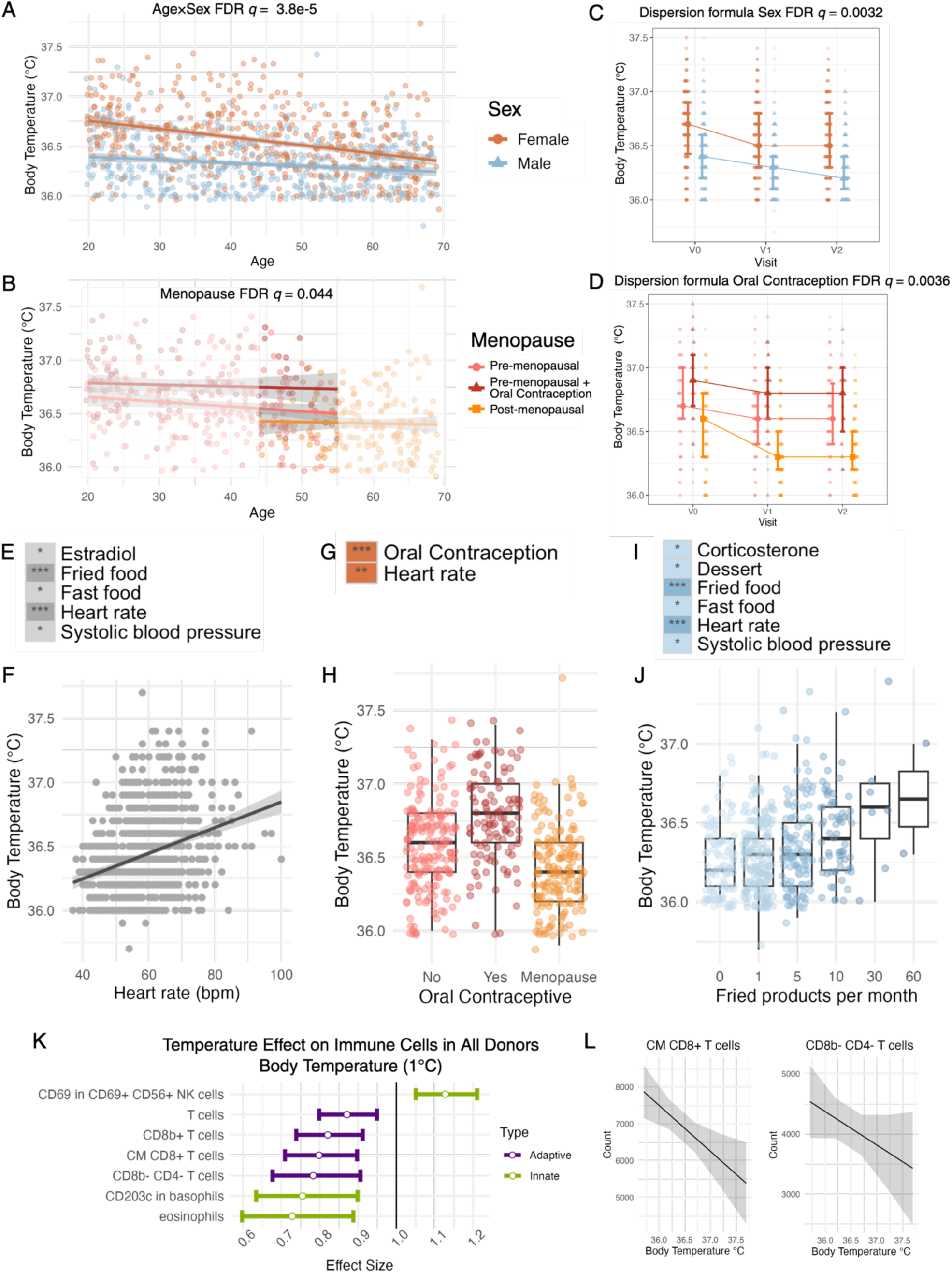
Body temperature variation in the *Milieu Intérieur* cohort. Scatter plots show MI V1 body temperature by age and applied regression models with the covariates of **A** age, sex and their interaction and **B** age and menopause in females 44-55 years old (with pre-menopausal females colored by oral contraception use for further illustration). **C** MI body temperature of 500 male and female donors over 3 visits spanning 8 weeks with applied mixed model and dispersion model. **D** MI body temperature of 250 female donors over 3 visits spanning 8 weeks with applied mixed model and dispersion model. Heatmap of significant associations between body temperature and 153 electronic case report from and steroid hormone variables identified by likelihood ratio tests of **E** all donors controlling for sex, age, and their interaction or controlling for age in **G** female donors and **I** male donors separately applying FDR correction. Values were plotted for the most significant novel association for each sub-cohort analysis: **F** heart rate in all donors (n=949), **H** oral contraceptive pill usage for female donors (n=472), and **J** fried food consumption for male donors (n=477), * FDR q value <0.05, ** <0.01, *** <0.001. **K** Significant effects (FDR *q <* 0.05) of body temperature on circulating adaptive or innate immune cells and their cell surface markers in all donors. Estimates derived from applied linear mixed models with log-transformed immunophenotype as the response, controlling for factors previously found to affect these parameters, then transformed to the original scale. Adaptive immune parameters in purple, innate immune parameters in green. **L** Partial effect plots of the most significant associations by body temperature.

To investigate which factors may predict body temperature in addition to age and sex, we analyzed 136 factors collected by questionnaire^26^ and plasma levels of 17 steroid hormones measured by mass spectrometry^27^ using likelihood ratio testing (LRT). To maximize statistical power, we first analyzed all donors together while controlling for age, sex, and their interaction. Of the 153 variables, we identified heart rate (β=0.0065) and fried foods eaten per month (β=0.0085) as most significantly associated with body temperature, along with systolic blood pressure (β=0.0022), number of fast-food meals per month (β=0.0059), and estradiol levels (β=-0.029) (**Figure 1E**). Heart rate was the only factor associated with body temperature in both sexes, displaying a positive correlation and emphasizing the inextricable link between these two homeostatic measures (**Figure 1E,F,G,I**)^28,29^.

Since the regression analysis of all donors revealed estradiol, a sex hormone, as associated with body temperature, we assessed whether these factors correlated with body temperature differently depending on sex (see Methods). When analyzing the sexes independently, oral contraception (β=0.18) and heart rate (β=0.0070) were positively correlated with female body temperature, consistent with previous studies (**Figure 1G,H**)^10^. Male body temperature was most significantly associated with heart rate (β=0.0062) and frequency of fried food consumption (β=0.0096), as well as systolic blood pressure (β=0.0032), levels of corticosterone (β=-0.042), and the frequency of eating dessert (β=0.0034), and fast food (β=0.0059) (**Figure 1I**).

The associations between body temperature and dietary habits were consistent across all donors and males (**Figure 1E,I)**, with fattier diets correlating with higher body temperature (**Figure 1J**). Steroid hormone effects emerged as sex-specific correlates of body temperature, with estradiol in the pooled analysis, corticosterone in males, and oral contraception use in females, highlighting the importance of sex-stratified analyses.

We next sought to determine how body temperature influences immune cell composition, by applying linear mixed models to flow cytometry data^30^, while controlling for sex, age, age×sex interaction, and additional covariates (see Methods). In the innate cell compartment, eosinophil counts were negatively correlated with body temperature along with cluster of differentiation (CD)203c expression in basophils (**Figure 1K**). Additionally, the activation marker CD69 on CD69^+^CD56^+^ natural killer (NK) cells was positively correlated with temperature, a relationship observed previously in T cells^21^. In the adaptive compartment, the counts of CD3^+^ T cells and three T cell subsets, CD8b^+^ T cells, central memory (CM) CD8b^+^ T cells, and CD8b^-^CD4^-^T cells, were all negatively correlated with body temperature. From calculated effect sizes, we estimated that a 1.5°C increase in body temperature correlated with a 0.5 log_2_ decrease in the amount of circulating CD8b^+^ T cells and CM CD8b^+^ T cells (**Figure 1L**). Together these results indicate a significant attenuation of baseline innate and adaptive immune function with increasing temperature.

### Sex-specific effects of homeostatic body temperature variation on immune responses

To characterize the effects of body temperature on immune responses, we fit multiple regression models to interrogate how MI donor body temperature was associated with differences in *ex vivo* induced immune responses. As previously described^4,31,32^, blood was stimulated *ex vivo* in TruCulture tubes for 22 hours at 37°C with a range of stimuli, and immune responses were characterized with transcriptomics and proteomics (**Figure 2A**, see Methods)^33^. We first analyzed the nonstimulated control condition, modeling the log_2_ gene expression with body temperature, while controlling for LRT-selected covariates and multiple testing (see Methods). Given the age×sex interaction in body temperature, we included sex and age×sex as covariates in the combined analysis as well as analyzed female and male donors independently.

**Figure 2.**
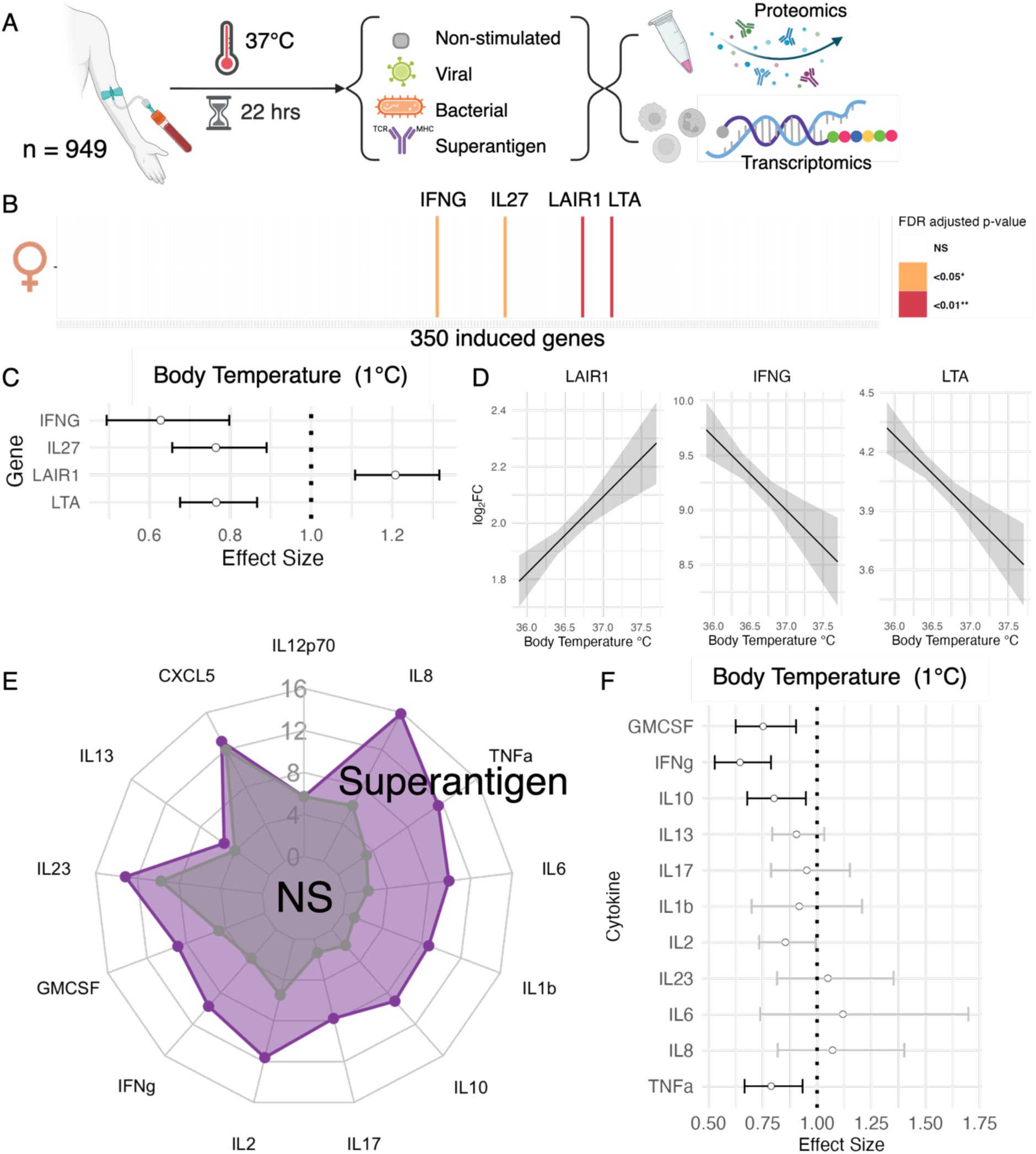
Body temperature effects on adaptive immune responses in female donors. **A** Experimental design **B** Barcode plot of genes induced by superantigen (SEB) stimulation (log_2_FC over nonstimulated >1) with those whose response is associated with female body temperature highlighted in color (FDR *q* < 0.05 in orange, *q* < 0.01 in red) **C** Quantification of the effect of body temperature on gene expression (log_2_FC over the nonstimulated control condition) post superantigen stimulation in female donors. These were estimated in a multiple regression model controlling for age, 16 major cell subtype proportions, periodic time and day of sampling, smoking status, and cytomegalovirus (CMV) serostatus then transformed to the original scale. Only genes with significant effect sizes (FDR corrected *q*-values < 0.05) were plotted. **D** Body temperature partial effect plots on the expression of the most significantly associated genes. **E** Spider plot showing cytokine levels in nonstimulated (grey) and superantigen (purple) condition for females; cytokines were considered induced if the levels in the stimulated condition were doubled from the nonstimulated control. **F** Quantification of the effect of body temperature on log2-transformed induced cytokine levels in the superantigen stimulation condition for females, effect sizes with FDR corrected *q*-values < 0.05 are in black, nonsignificant effects in grey (n=472)

In the nonstimulated condition, six genes were associated (false discovery rate (FDR) *q*<0.05) with body temperature in all donors, three of the same genes when analyzing female body temperature, and two genes unique to males (**Supp Fig 1A**). In all donors, the expression of *SH2D1A*, *GZMK*, *LAG3*, *SLAMF1*, and *CD1C0* decreased with higher body temperature, whereas *TLR2* expression increased (**Supp Fig 1B**). Females showed the same negative correlations for adaptive immune regulators *LAG3*, *SH2D1A*, and *SLAMF1*, with larger effect sizes and confidence intervals **(Supp Fig 1C**). Males uniquely showed positive correlations between body temperature and the type I IFN signaling genes *MX1* and *STAT2* (**Supp Fig 1D**). Given these baseline sex-specific associations, and established sex differences in MI immune responses^4,33^, we kept a sex-stratified approach in subsequent analyses of stimulated conditions.

We extended this analysis to genes and cytokines induced (log_2_ fold change (FC) > 1 over nonstimulated condition) by superantigen (staphylococcal enterotoxin B (SEB); T cell agonist), viral (polyinosinic:polycytidylic acid (Poly I:C); TLR3, retinoic acid inducible protein I (RIG-I), melanoma differentiation-associate gene 5 (MDA-5) agonist), and bacterial (*E. coli*; TLR2/4 agonist) stimulation conditions (see Methods). In female donors, body temperature was uniquely associated with immune responses to superantigen stimulation (**Figure 2**, **Supp Fig 2B,C**). Of the 305 genes induced after superantigen stimulation, the transcriptional response of *LAIR1*, *LTA*, *IFNG*, and *IL27* were associated with body temperature (**Figure 2B**). A 1°C increase in body temperature was predicted to decrease pro-inflammatory *IFNG* response by 37% on average and increase inhibitory *LAIR1* response by 20% (**Figure 2C, D**). At the proteomic level, more than one third of the superantigen-induced cytokines in females were negatively correlated with body temperature: IFN-γ, tumor necrosis factor alpha (TNFα), granulocyte-macrophage colony-stimulating factor (GM-CSF), and IL-10 (**Figure 2E**). As was observed at the transcriptomic level, we confirmed that 1°C increase in body temperature decreased IFN-γ by 36% on average (**Figure 2F**).

In contrast to female donors, male body temperature was specifically associated with transcriptional responses upon viral and bacterial stimulation (**Figure 3, Supp Fig 2 A**). In the viral condition, 24% of the induced genes (36/150) were associated with body temperature. The genes most significantly impacted by body temperature were *MX1*, *IFIT2*, *STAT2*, *TNFSF10*, and *TNFSF13B* (**Figure 3A**). The estimated effect of a 1°C change in body temperature ranged from a 9% to 45% change in transcriptional response (**Figure 3B**). Notably, 33 out of 36 temperature-associated genes were up-regulated with viral stimulation and exhibited weaker induction at high temperatures, while the remaining three genes were down-regulated with stimulation and displayed weaker repression at high temperatures, suggesting a dampening of anti-viral immune responses with rising temperatures (**Figure 3C** and **Supp Table 1**). A similar pattern was observed with the cytokine data: two of the highest-induced cytokines, IL-6 and IL-12p70 were negatively correlated with body temperature (**Figure 3D-F**). On average, a 1°C increase in body temperature decreased IL-6 levels by 27% and IL-12p70 levels by 35% (**Figure 3E**).

**Figure 3.**
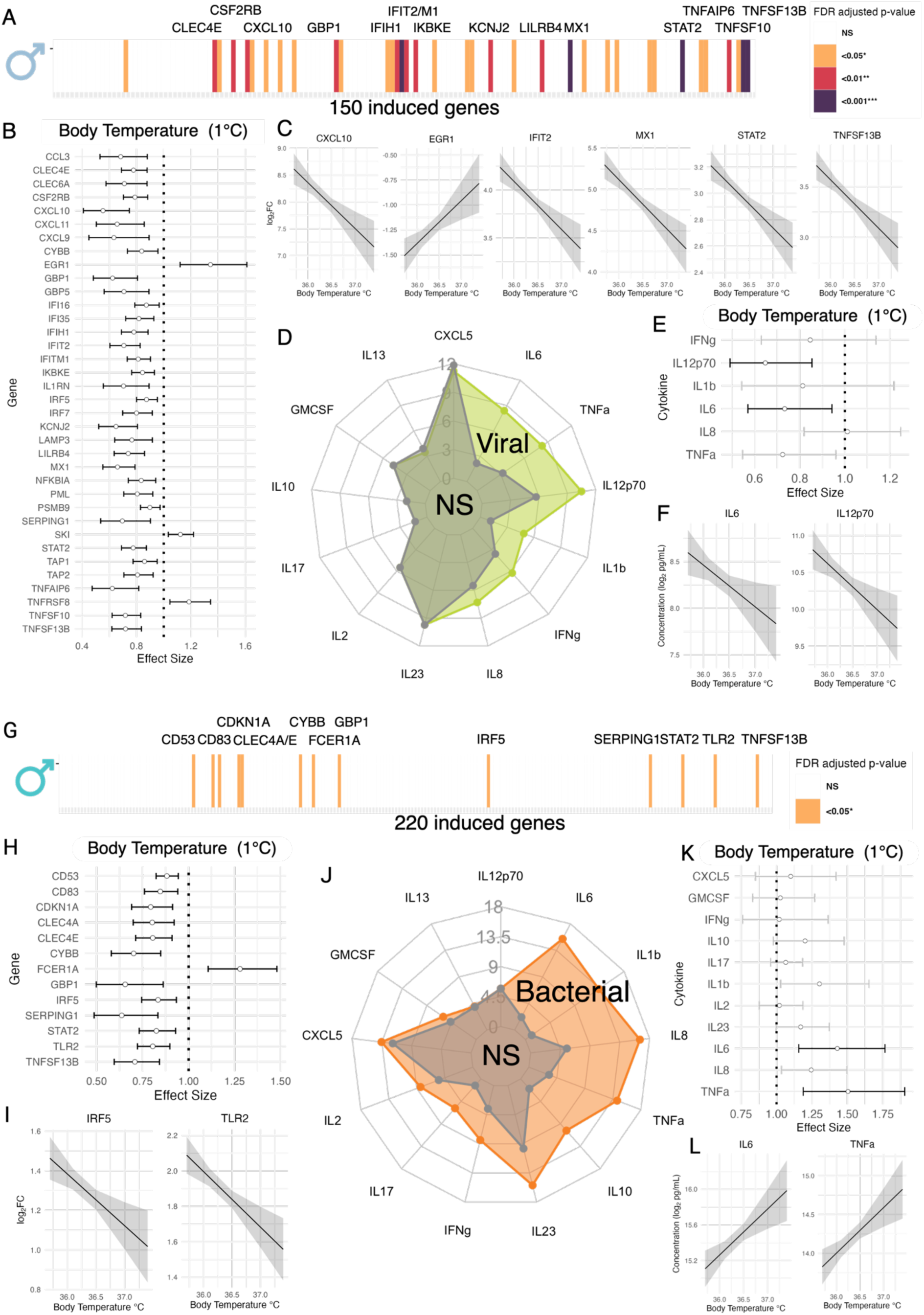
Body temperature effects on the immune response to viral stimulation in male donors. Barcode plot of genes induced by **A** Viral (Poly I:C; TLR3, RIG-I, MDA-5 agonist), or **G** Bacterial (*E. coli*; TLR2/4 agonist), stimulation (log_2_FC over nonstimulated >1) with those whose expression is associated with male body temperature highlighted in color (FDR *q* < 0.05 in orange, *q* < 0.01 in red, *q* < 0.001 in purple). **B** Viral, **H** Bacterial quantification of the effect of body temperature on gene expression (log_2_FC over the nonstimulated control condition) post stimulation in male donors. These were estimated in a multiple regression model controlling for age, 16 major cell subtype proportions, periodic time and day of sampling, and smoking status then transformed to the original scale. Only genes with significant effect sizes (FDR corrected *q*-values < 0.05) were plotted. **C** Viral**, I** Bacterial body temperature partial effect plots on the expression of the most significantly associated genes. **D** Viral (green), **J** Bacterial (orange) spider plot showing cytokine levels in nonstimulated (grey) and stimulated (color) conditions in males; cytokines were considered induced if the levels in the stimulated condition were doubled from the nonstimulated control. **E** Viral**, K** Bacterial quantification of the effect of body temperature on log2-transformed induced cytokine levels in the stimulation condition for males, effect sizes with FDR corrected *q*-values < 0.05 are in black, nonsignificant effects in grey. **F** Viral**, L** Bacterial body temperature partial effect plots on the levels of significantly associated cytokines. (n=477)

In the bacterial condition, the response of 6% of the induced genes (13/220) were associated with body temperature, including seven which overlapped with the viral findings: *CLEC4E*, *CYBB*, *GBP1, IRF5, SERPING1*, *STAT2,* and *TNFSF13B* (**Figure 3G**). For 12 of these genes, transcriptional response decreased by 12% to 36% per 1°C increase in body temperature and only one gene (*FCER1A*) had increased response with increased temperature (**Figure 3H**). As with viral stimulation, the gene positively correlated with body temperature (*FCER1A*) was down-regulated after bacterial stimulation, while the genes that were negatively correlated with temperature (e.g. *IRF5* and *TLR2*) were up-regulated (**Figure 3I** and **Supp Table 1**). The levels of the highest-induced cytokine, IL-6, and TNFα, were positively correlated with body temperature (**Figure 3J-L**). On average, a 1°C increase in body temperature increased IL-6 levels by 57% and TNFα levels by 50% after bacterial stimulation (**Figure 3K**).

### Febrile temperature effects on healthy immune responses *ex vivo*

Beyond homeostatic variation, changes in body temperature are notably observed with fever, a hallmark symptom of disease. After characterizing immune responses to temperature change within the healthy range, we investigated how these responses may change under febrile temperatures by performing TruCulture stimulations on healthy donor whole blood (n=20 donors, 10 female, 10 male) for 22 hours at homeostatic (36.8°C) and febrile (39°C) temperatures (**Figure 4A)**. Once again stimulation conditions inducing viral (resiquimod (R848); TLR7/8 agonist), bacterial (lipopolysaccharides (LPS) component of *E. coli,* TLR4 agonist), and superantigen (CytoStim) inflammatory pathways were compared against a nonstimulated control, and immune responses were characterized with transcriptomics and proteomics. Linear mixed models including a stimuli×temperature interaction were fit to induced cytokines to assess stimulation-dependent effects of temperature (see Methods).

**Figure 4.**
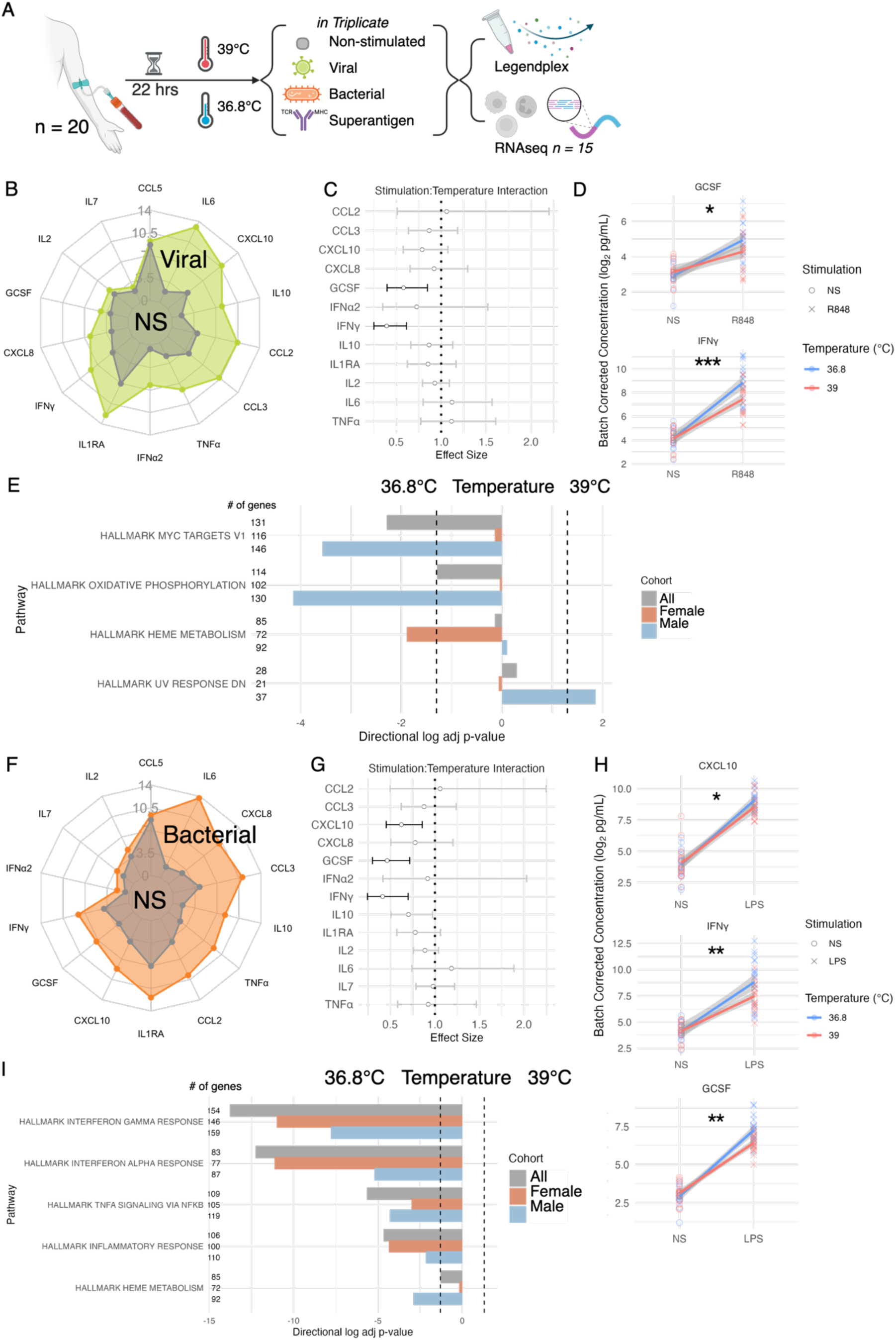
Febrile temperature reduces cytokine responses in *ex vivo* whole blood stimulations. **A** Experimental design **B, F** Spider plots showing cytokine levels in the supernatant of healthy whole blood stimulations (n = 20) after 22 hours of incubation in the nonstimulated (grey) or stimulation (viral, R848= green, bacterial, LPS = orange) condition; cytokines were considered induced if the levels in the stimulation condition were doubled from the nonstimulated control. **C, G** Quantification of the interaction effect of stimulation and incubation temperature (36.8°C or 39°C), on the abundance of induced cytokines estimated using a linear mixed model with log_2_-transformed concentration as the response, controlled for donor and batch effect, then transformed to the original scale. Effect sizes with FDR corrected *q*-values < 0.05 are in black, nonsignificant effects in grey. **D, H** Models that showed a significant interaction between temperature and stimulation were visualized with line graphs of batch-adjusted cytokine concentrations. **E, I** Differentially expressed Hallmark gene sets significantly downregulated in cells from a subset of the viral (R848) or bacterial (LPS) stimulated healthy whole blood samples (n=15) at 39°C vs 36.8°C

Of the 12 cytokines induced by viral stimulation (**Figure 4B**), incubation temperature influenced the expression of IFN-γ and granulocyte colony stimulating factor (G-CSF), as evidenced by a stimulation×temperature interaction (**Figure 4C**). Estimated effects showed febrile temperature reduced IFN-γ levels by 60% and G-CSF levels by 42% (**Figure 4C**). These attenuated cytokine levels are particularly evident when plotting batch-corrected log2 concentrations (**Figure 4D**). Of the 13 cytokines induced by bacterial stimulation (**Figure 4F)**, febrile temperature again correlated with reduced levels of IFN-γ, G-CSF, and chemokine ligand 10 (CXCL10), with an estimated effect of up to 39% for IFN-γ (**Figure 4G**). Residual plots illustrate this relationship between increased incubation temperature and attenuated cytokine levels (**Figure 4H**). There was no significance after FDR correction in the superantigen condition or with temperature alone in the nonstimulated control (**Supp. Fig. 3A**).

To explore coordinated transcriptomic temperature effects, we performed gene set tests on RNAseq data (see Methods) from a subcohort stratified by sex (n=11 donors, 6 female, 5 male). After viral stimulation, we found febrile temperatures downregulated Oxidative Phosphorylation and MYC Targets for all donors and males, and Heme Metabolism in females. After bacterial stimulation, febrile temperatures downregulated induced IFN-α and IFN-γ Response, TNFα Signaling via NF-kB, and Inflammatory Response across all subgroups, demonstrating pleiotropic regulatory effects on bacterial immunity (**Figure 4G**).

By manipulating temperature directly within the same donor samples, these experiments establish elevated temperature as an independent regulator of immune responses, with febrile temperatures sufficient to attenuate key antiviral and antibacterial pathways.

### Cell-specific temperature effects

To identify which immune cell subsets underlie the observed sex-specific temperature effects, we performed additional *ex vivo* stimulations (superantigen CytoStim, viral Poly I:C, bacterial *E.coli*, nonstimulated control) at homeostatic (36.8°C – 37°C) and febrile temperatures (39°C) coupled with cellular indexing of transcriptome and epitope sequencing (CITE-seq)^32^(n=6, 3 female, 3 male). (**Figure 5A**, see Methods). After a 4-hour whole blood stimulation, we isolated peripheral blood mononuclear cells (PBMCs), labeled them with 20 antibodies, and captured over 950,000 high-quality single-cell transcriptomes. Using transcriptome-based harmonized clusters (**Supp Fig 4A**), CITE-seq marker expression **(Supp Fig 4C)**, and a cell-assignment model previously trained on 1.5 million stimulated PBMCs, we identified 42 cell types across myeloid, B, T, and NK lineages (**Figure 5B, Supp Fig 4B**). Cell types with the greatest sex bias were CD56dim NK cells, which showed a 55% relative increase in males compared to females (males:14.6%, females:9.4%), while mucosal-associated invariant T cells (MAIT) cell proportions were doubled in females (females:5.5%, males:2.7%)(**Figure 5C**), consistent with MI data^30^. CD14^+^ classical monocyte proportions were similar at homeostatic temperatures (females:11.2%, males:11.5%), but females showed a 20% relative increase at 39°C (homeostatic:11.2%, febrile:14.0%), while males showed no change (homeostatic:11.5%, febrile:11.7%), indicating potential sex-specific temperature effects on CD14 expression.

**Figure 5.**
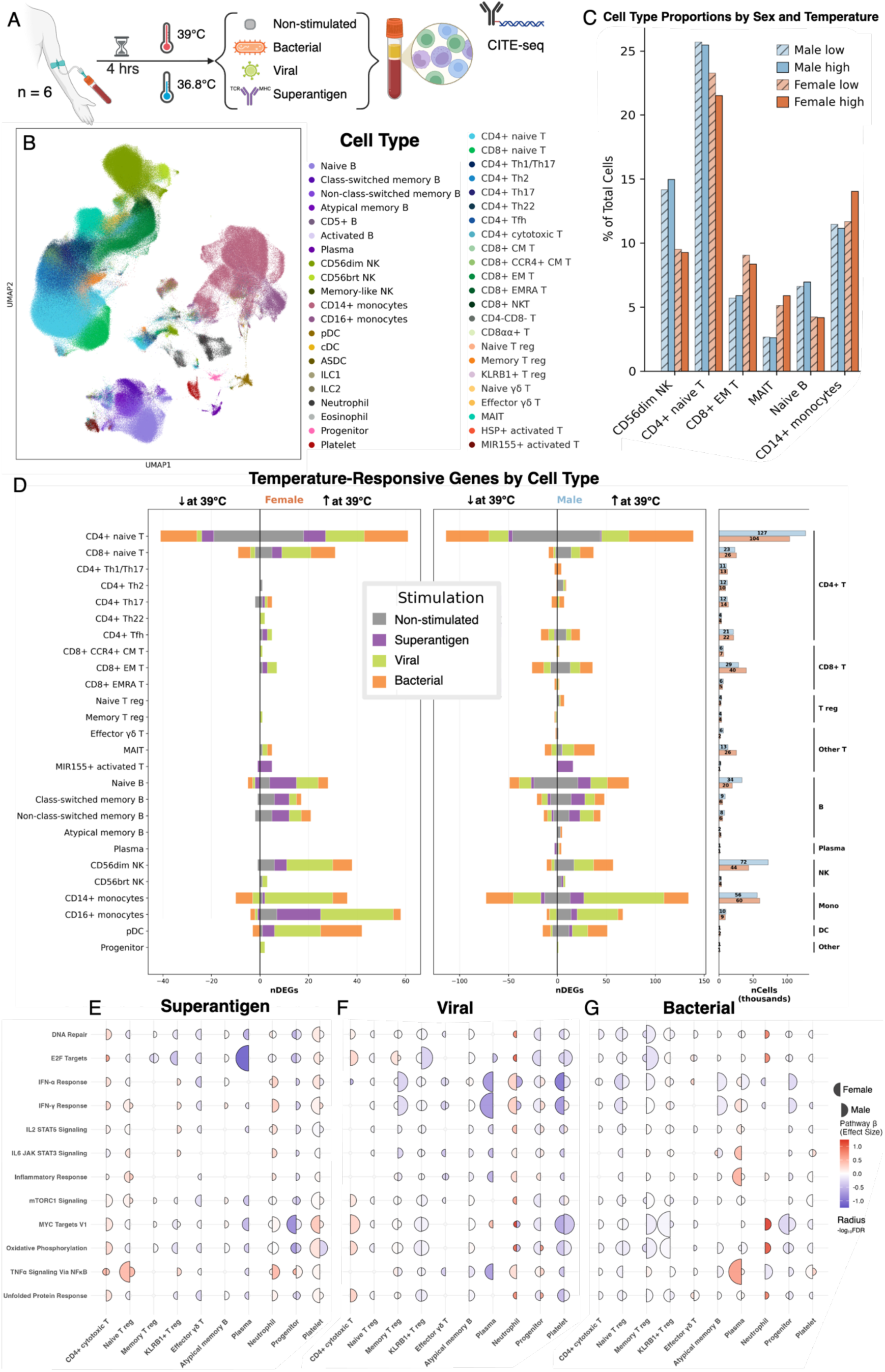
Single-cell resolution of temperature-induced immune responses reveals sex-specific immune adaptations. **A** Experimental design: Whole blood from healthy donors (n=6) was cultured in TruCulture tubes at physiological (36.8°C) or febrile (39°C) temperatures for 4 hours under four conditions: non-stimulated control, superantigen (CytoStim; T cell activation), viral (Poly I:C; TLR3, RIG-I, MDA-5 agonist), bacterial (*E. coli*; TLR2/4 agonist), followed by PBMC isolation and CITE-seq analysis. **B** Stimulation-harmonized UMAP visualization of 42 immune cell types identified across all conditions. **C** Cell type proportions stratified by biological sex and temperature, displaying the top 6 cell types with sex differences. **D** Barplot quantifying temperature-responsive genes (39°C vs 36.8-37°C) across cell types and stimulation conditions (grey = negative control, purple = superantigen, green = viral, orange = bacterial). Hallmark gene set test analysis showing the top 12 temperature-responsive gene sets and top 10 cells with sex differences in pathway response across each stimulation condition: **E** superantigen, **F** viral, and **G** bacterial. Dot size indicates −log10(FDR) and color represents direction and strength of effect across the gene set (red = upregulated, blue = downregulated at 39°C vs 36.8°C). Female dots are the left hemisphere and male dots are the right hemisphere.

Thirty-one cell types showed transcriptional changes with temperature in at least one stimulation condition (**Figure 5D, Supp Fig 4D**). Naïve CD4+ T cells had the most temperature DEGs and were also the most abundant cell type. plasmacytoid dendritic cells (pDCs) were the most temperature responsive cell type when normalizing DEGs per 1000 cells. Heat shock proteins were predictably upregulated at 39°C, while immune-regulatory genes such as *HLA-F* were downregulated (**Supp Fig 4E)**. Superantigen stimulation accounted for a larger proportion of female temperature DEGs (18.8% vs 8.0%), and bacterial stimulation a larger proportion of male temperature DEGs (33.5% vs 24.3%).

Sex-stratified gene set analysis revealed coordinated temperature effects on immune pathways, with females having more temperature responsive gene sets in the negative control condition and in the T cell compartment following superantigen stimulation, while in males more temperature responsive gene sets were present after viral and bacterial stimulations (**Supp Fig 5B**). Prioritizing the most temperature-responsive gene sets and cell types with the greatest sex differences in response (**Figure 5E-G**), TNFα Signaling via NF-kB and IFN-γ Response were downregulated by temperature during superantigen stimulation in female effector γδ T and plasma cells, but upregulated in more protective naïve T regs and platelets (**Figure 5E**). Temperature downregulated cell growth and immune signaling signatures in superantigen-stimulated female pDCs, neutrophils, and progenitor cells, whereas these signatures were upregulated in males (**Supp Fig 5B**).

With viral stimulation, higher temperatures downregulated Inflammatory Response, IFN-α and IFN-γ Responses, or TNFα Signaling via NF-kB across 7 of the 10 cell types with the greatest sex difference in temperature response (**Figure 5F**), consistent with our earlier bulk findings (**Figures 3, 4)**. In males, these gene sets were most strongly attenuated in neutrophils, which in contrast were enhanced in females (**Figure 5F**). Bacterial stimulation showed similar patterns, as higher temperatures downregulated IFN-α and IFN-γ responses across 22 and 24 cell types in males respectively, versus 14 and 16 cell types in females (**Supp Fig 5B**). These effects are consistent with the bacterial response trends we saw at homeostatic temperatures (**Figure 3)** and mirror the bulk bacterial experiments (**Figure 4**) while providing single-cell resolution to these observations. KLRB1+ T regs were the only cell type in which IFN-α and IFN-γ Responses were upregulated by higher temperatures in males, but downregulated in females, which show signs of mitochondrial dysfunction reminiscent of HSR^35^. Among the 7 cell types where IFN-α and IFN-γ Responses were downregulated in males alone, CD56dim NK cells were notable given their two-fold greater abundance in males (**Supp Fig 5B**). These findings collectively suggest that sex differences in cell type abundance and temperature response may jointly drive immune response differences between females and males.

### Febrile temperature effects on SARS-CoV-2 immune responses *in vivo*

To determine whether the temperature-dependent immune effects identified in healthy donors persisted during active viral infection *in vivo*, we examined whole blood transcriptional responses in relation to body temperature in a clinical cohort of 82 COVID-19 patients (26 females, 56 males; **Figure 6A, 6B**). At the time of sampling these patients were already exposed to SARS-CoV-2 virus (TLR3/7, MDA5), making the TruCulture nonstimulated control condition a readout of *in vivo*-primed immune responses, and a physiologically relevant complement to our *ex vivo* viral stimulation models. Differentially expressed genes were identified in relation to both continuous body temperature and binary fever status (≥37.5°C), controlling for age, sex, disease severity, and periodic time of sampling. Temperature-associated transcriptional responses were detected in the full cohort (**Supp Fig 6A**) and in male patients (n=56; **Figure 6C**), but not in female patients (n=26; **Supp Fig 6C-D**), likely due to the limited statistical power at n=26.

**Figure 6.**
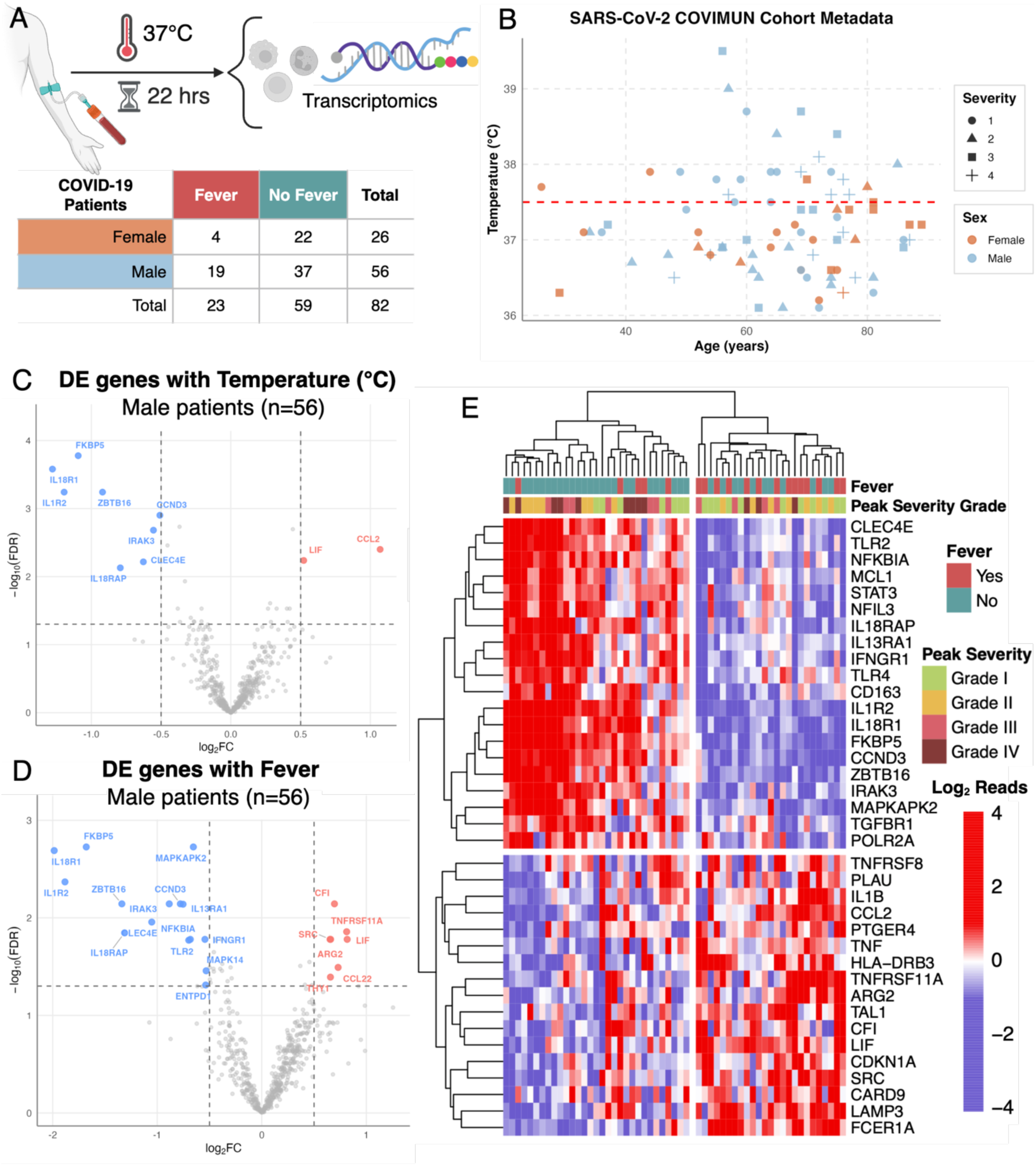
Temperature effects in SARS CoV-2 Infection. **A** Experimental design and table of cohort by sex and fever status. **B** Scatter plot of patient metadata showing age on the x-axis, body temperature on the y-axis, sex by color (females in red, males in blue) and severity by shape (1 least severe = circle, 2 = triangle, 3 = square, 4 most severe = cross). Volcano plots of differentially expressed genes associated with **C** temperature and **D** fever male patients (n=56) **E** Heatmap of significant DE genes associated with fever in male patients (n=56) annotated with fever status (fever = red, no fever = blue) and peak disease severity (1 least severe = green, 2 = yellow, 3 = pink, 4 most severe = brown).

Analyzing continuous body temperature, increased temperature correlated with downregulation of innate immune regulatory genes in the full cohort (**Supp Fig 6B)** and male patients (**Figure 6C**). IL-18 receptor subunits *IL18R1* and *IL18RAP* were both downregulated, indicating reduced IL-18 sensing capacity at elevated temperatures. *IL1R2*, the decoy IL-1 receptor, was also downregulated in patients with higher body temperatures, consistent with our finding of reduced *IL1RN* response with higher homeostatic temperature in the MI male viral condition (**Figure 3**). *IRAK3*, a negative regulator of TLR and IL-1R signaling, was also downregulated, as was *CLEC4E*, an innate pattern recognition receptor previously identified as temperature-sensitive in the MI male donors (**Figure 3**). Additional downregulated genes included *FKBP5*, consistent with HSP stabilization of glucocorticoid signaling as a potential target of temperature-associated modulation during active infection.

Binary fever analysis revealed an expanded and mechanistically coherent set of temperature-associated genes in the full cohort (**Supp Fig 6B**) and male patients (**Figure 6D**). Fever was associated with downregulation of *TLR2*, previously shown to directly sense the SARS-CoV-2 envelope protein^36^. This finding mirrors the association between the downregulation of *IFIH1*, a cytosolic sensor of SARS-CoV-2, with higher homeostatic temperatures in MI males (**Figure 3**), collectively indicating that elevated body temperature correlates with attenuated innate viral nucleic acid recognition.

Downstream of receptor-level sensing, fever was associated with downregulation of *MAPK14* and its substrate *MAPKAPK2*. Their axis post-transcriptionally stabilizes AU-rich element-containing inflammatory mRNA transcripts^37^, such as *IL-C*, *IL-12*, and *TNFα*, its attenuation provides mechanistic support for the reduced IL-6 and IL-12p70 levels observed with elevated temperature in MI males (**Figure 3**), and for the reduced IFN-γ production at febrile temperatures in *ex vivo* viral stimulation (**Figure 4**). Consistent with this upstream signaling attenuation, *NFKBIA* was downregulated with fever in SARS-CoV-2 infection, paralleling its negative association with higher homeostatic temperature in the MI male cohort (**Figure 3**). The simultaneous downregulation of *NFKBIA*, *MAPK14*, and *IRAK3* indicates attenuation of nuclear factor kappa light chain enhancer of activated B cells (NF-κB) and mitogen activated protein kinase (MAPK) signaling as shared upstream mechanisms underlying temperature-associated reductions in IL-6, IL-12p70, and IFN-γ observed across all three cohorts.

Attenuation of IFN-γ responses was further evidenced by the negative correlation between *IFNGR1* expression with fever in male patients, extending attenuation beyond IFN-γ production to cellular responsiveness to this cytokine. The IL-18 signaling axis was again well represented: *IL18R1*, *IL18RAP*, and *IRAK3* were downregulated in febrile patients, consistent with the continuous temperature findings. LILR family downregulation was also observed: *LILRB3* in all febrile COVID-19 patients, extending the *LILRB4* finding in MI males (**Figure 3**), along with *IL1R2* and *CLEC4E* were again downregulated in febrile patients, reinforcing them as temperature-associated targets across experimental and disease contexts.

To determine whether these gene-level differences reflect broader, coordinated immune states, we performed unsupervised hierarchical clustering of DEGs across individual male patients (**Figure 6E**). The heatmap revealed two patient clusters that were strongly associated with fever status. The gene block elevated in non-febrile patients was enriched for pattern-recognition receptors, NF-κB negative regulators, and anti-inflammatory signaling molecules, suggesting a controlled innate antiviral response. The gene block elevated in febrile patients contained canonical pyrogenic cytokines, monocyte chemoattractants, antigen-presentation machinery, and complement/coagulation-related factors, consistent with progression to systemic hyperinflammation.

### Multicellular analyses link febrile temperature to suppressed male antiviral responses

To identify candidate cellular mechanisms underlying the male-specific temperature effects on viral responses observed in healthy donors (**Figure 3**) and COVID-19 patients (**Figure 6**), we reconstructed multicellular regulatory networks from the 4h viral stimulation single-cell data using ReCoN (REconstruction of multicellular COordination Networks from single-cell data)^38^ (see Methods). ReCoN and complementary transcriptomic analyses identified class-switched memory B cells, AXL+ SIGLEC6+ dendritic cells (ASDCs), and CD14+ monocytes as the primary sender populations driving temperature-modulated responses, with CD56dim NK cells, CD4+ Tfh cells and atypical memory B cells as the primary receiver populations (**Figure 7A**, see Methods).

**Figure 7.**
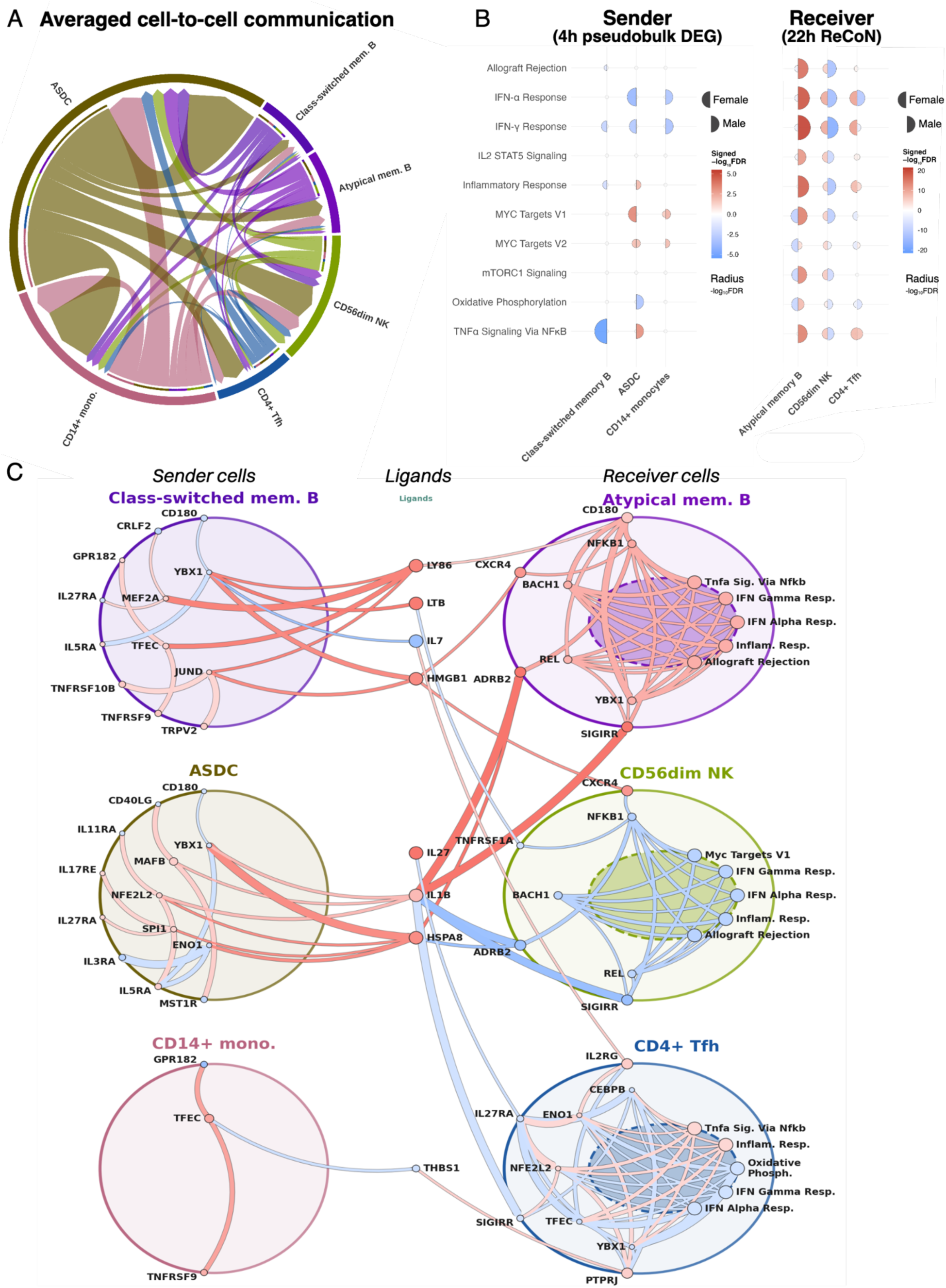
Temperature regulation of male viral responseA. Chord diagram of averaged cell-to-cell communication between temperature-responsive cell types in males across temperatures after 4h of viral stimulation **B** Hallmark gene set tests showing the top temperature-responsive gene sets in the viral condition. **Left** Sender cells’ pathway results from 4h pseudobulk counts **Right** Receiver cells’ pathway results from downstream ReCoN scores. Dot size and color indicate −log10(FDR) and direction of regulation (red = upregulated, blue = downregulated at 39°C vs 36.8°C). Female dots are the left hemisphere, and male dots are the right hemisphere. **C** Pseudotime cascade plots for the male viral stimulation condition showing temperature-modulated cell-to-cell communication. The left column shows the 3 “sender cells”, the middle column their differentially expressed molecular signals being sent to the receiver cells in the right column. Dots on the sender cell membranes represent the top receptors through which they are being regulated, signaling to the cytoplasm dots representing the top transcription factors modulating differentially expressed ligand genes shown in the column between the sender cells and receiver cell. These ligands are recognized by the receptor dots on the receiver cell membrane, signaling to the transcription factors represented by dots in the cytoplasm, and modulating gene expression in most significantly modulated Hallmark gene sets for that cell. Dot and connector color are determined by the temperature effect on gene expression or ReCoN score (red = upregulated, blue = downregulated at 39°C vs 36.8°C).

ReCoN predicted IFN-α and IFN-γ receiver responses would be the most significantly temperature-modulated gene sets (**Figure 7B**), consistent with our 22h data (**Figure 4)**. These responses were regulated in opposite directions for males and females in memory T regs, atypical memory B cells, CD56dim NK cells, CD14+ monocytes, and T follicular helper cells (**Figure 7B and Supp. Fig. 7**). Network analysis identified possible ligand-receptor axes regulating these modulations (**Figure 7C**). IL-1B, TNFRSF, and SIGIRR signaling from sender cells converged on CD56dim NK cells, downregulating IFN-α, IFN-γ, Inflammatory, and MYC Target hallmarks through NFκB and HSP-associated transcription factors^39^. Males displayed a 55% relative expansion in this NK population compared to females (**Figure 5C**), suggesting that temperature-dependent functional attenuation is compounded by greater abundance of this subset.

CD4+ Tfh cells, essential for mucosal and B cell immunity in COVID-19 responses^40^, showed downregulated Oxidative Phosphorylation alongside IFN-α and IFN-γ Response signaling, suggesting immunometabolic mediation of these temperature-immune effects through ENO1 and NFE2L2 (**Figure 7C**). Conversely, Memory T and B cells were the only subsets with enhanced IFN-α and IFN-γ Responses, including atypical memory B cells whose expansion is associated with severe COVID-19 and chronic inflammatory states ^41–43^. IL1B, SIGIRR, and NFKB1 signaling were again implicated in these cascades. Together, these results connect systemic clinical observations to defined multicellular circuits, and suggest that temperature tunes immune responses in a sex-specific manner through coordinated signaling and metabolic programs.

## Discussion

Body temperature is a key homeostatic parameter, yet the extent of its healthy variability and its influence on immune responses remains poorly defined, with a notable lack of population-level studies. Addressing these questions is critical for understanding the biological mechanisms underlying sex differences in immune function and disease prevalence. We conducted a multi-dimensional analysis of the impact of body temperature on immune responses in 975 healthy donors and 82 COVID-19 patients, revealing sex-specific effects: female immune responses to superantigen stimuli were attenuated with increased temperature, whereas male immune responses to bacterial and viral stimuli were suppressed. To our knowledge, this is the first large cohort study examining how healthy body temperature variation affects human immunity. Our findings suggest that body temperature is not only a consequence of immune activation, but also a physiological pressure that shapes immune responses and contributes to individual immune setpoints.

Females were on average 0.5°C warmer and showed greater temperature variation than males, consistent with previous studies^11,44^. Even within this narrow homeostatic range, warmer temperature was associated with circulating innate and adaptive immune cell proportions, including decreases in CD8^+^ and double-negative T cells. In addition to previously described^30^ sex biases in the proportion of NK cells for males, and MAIT and naïve CD8^+^ T cells for females, there was a 20% relative increase in female CD14^+^ classical monocytes in our *ex vivo* fever model, highlighting that temperature context is crucial for understanding sex differences in immune setpoints^45,46^.

Temperature was also associated with sex-specific functional differences in immune responsiveness across both homeostatic and febrile ranges. In female donors, higher body temperature was associated with attenuated superantigen-induced T cell responses. IFN-γ was negatively correlated with body temperature at transcriptomic and proteomic levels, along with the pyrogenic cytokine TNFα. Our findings suggest that elevated temperature, a classical hallmark of inflammation, may act as an anti-inflammatory signal in a negative feedback mechanism, analogous to recently characterized NOD like receptor protein 3 (NLRP3) inflammasome regulation^47^. This effect was most pronounced in double-negative, effector γδ, and CD8+ central memory T cells after superantigen-stimulation at febrile temperatures, with these adaptive populations showing four-fold more temperature-responsive hallmark gene sets than their male counterparts.

In male donors, temperature was associated with innate-obligate responses to bacterial and viral stimuli. Seven proinflammatory genes were negatively correlated with body temperature across both conditions, including genes involved in pathogen recognition, IFN-α induction, and oxidative stress responses like HSR, consistent with feedback inhibition of pyrogenic pathways as temperature rises^25^. Higher temperature was associated with lower pyrogenic IL-6 cytokine levels after viral stimulation. Febrile temperatures attenuated IFN-α and IFN-γ responses to a greater extent in males after 4 hours of viral and bacterial stimulation, with downregulation in nearly three times as many cell types. After 22 hours, this attenuation effect was mirrored by reduced levels of pro-inflammatory cytokines including IFN-γ, G-CSF and CXCL10. Gene set testing confirmed this modulation at a transcriptomic level, suggesting alternative pathways through which hyperthermia modulates immune responses, potentially through metabolic or steroidal intermediates^25^.

Clinical validation in COVID-19 patients confirmed that fever-range temperatures attenuate anti-viral responses in males through conserved IL-1β, NF-κB, IFN-γ, and MAPK pathways. Multi-layered single-cell regulatory modeling traced these effects to class-switched memory B cells, ASDCs, and CD14+ monocytes, which engaged atypical memory B cells, CD56dim NK cells, and CD4+ Tfh cells via IL-1β, HSPs, IL-27, and IL-7 signaling. Oxidative phosphorylation emerged as a metabolic checkpoint, supporting a mechanistic role for immunometabolism in sex-specific immune modulation^48^. Our three complementary study systems together provide compelling rationale for integrating sex and body temperature into precision medicine frameworks for antiviral treatment and managing chronic inflammatory states.

Across cohorts and experiments, temperature responses were consistently greater in the female adaptive immune compartment and the male innate immune compartment. These patterns mirror sex disparity in disease susceptibility, with males more susceptible to pathogen-associated damage in infectious disease and females more prone to autoimmune disorders driven by pathogenic T cell activity^3^. Life history theory proposes sex-specific reproductive roles and resource allocation has driven greater investment in innate immunity for males and adaptive immunity for females^49–52^. We hypothesize further that temperature acts as a fine-tuning mechanism on these compartments, and may contribute to sex differences in immune setpoints and disease risk, and could even be exploited for therapeutic effects.

Our study has limitations. MI limited diversity by design to diminish confounding factors in immune variability, therefore our research should be replicated in diverse cohorts to determine whether our findings are universal or vary with genetic, endocrine, and cultural factors. MI *ex vivo* stimulations were performed uniformly at 37°C, yet body temperature associations with induced responses remained detectable, indicating a lasting effect of this homeostatic parameter on personal immune setpoints. The febrile experiments and COVID-19 cohort had limited sample sizes, particularly for females, and future sex-balanced clinical studies and larger single-cell datasets could more confidently identify sex differences in immune responses at febrile temperatures. Integrating menstrual cycle data into longitudinal sampling would help clarify endocrine-temperature-immune interactions. Finally, our study focuses on heat, while cold also modulates immunity, as seen in cryopyrin-associated periodic syndromes and accidental hypothermia^53,54^, and warrants systems level investigations.

In conclusion, natural variability in body temperature attenuates immune responses across homeostatic and febrile ranges and has sex-specific consequences. Understanding the complex mechanisms underlying homeostatic interactions between body temperature, metabolism, and immune responses provides insight into the sources of population-level immune variation and can pave the way for guided personalized interventions in the face of escalating extreme heat events^55,56^. Harnessing temperature effects on immunity could lead to new therapies, as already illustrated by hyperthermia as an adjuvant for cancer treatments, an anti-inflammatory treatment for ankylosing spondylitis, and a microwave-induced solution for refractory human papillomavirus skin lesions^20,21,57–59^.

## Materials and Methods

### Milieu Interieur Clinical Protocol

Human samples were collected as part of the *Milieu Interieur* cohort, which was approved by the *Comité de Protection des Personnes—Ouest C* and the *Agence Nationale de Sécurité du Médicament* in June 2012 and sponsored by the Institut Pasteur (ID-RCB no. 2012-A00238-35). The study protocol was designed and conducted in accordance with the Declaration of Helsinki and good clinical practice as outlined in the International Conference on Harmonisation of Technical Requirements for Registration of Pharmaceuticals for Human Use (ICH) Guidelines for Good Clinical Practice. The original protocol was registered under ClinicalTrials.gov (NCT01699893). All donors gave written informed consent.

The cohort consists of 949 healthy donors selected based on inclusion and exclusion criteria that were previously described^60^. Briefly, donors had no history or evidence of pathological conditions, substance abuse, or recent vaccination. Donors are equally distributed amongst males and females across 5 decades of life (20 – 69 years old). To decrease potential confounding variability, all donors self-declared to have 3 generations metropolitan French ancestry, and were not pregnant or perimenopausal. Sampling took place in the morning at a single site (Biotrial, Rennes 35000) over a continuous 11-month period from September 2011 to August 2012.

#### Datasets

##### Case Report Form variables

Lifestyle, dietary, and medical history factors were collected in an electronic case report form based questionnaire as previously described^60^. Clinical parameters such as temperature and blood pressure were measured on the day of blood sampling by Biotrial staff and appended to the electronic case report form. To select factors for analysis, we selected variables that had data for at least 5% of individuals in at least two categorical levels.

##### Steroid Hormones

To obtain free and non-covalently bonded steroid hormone measures, plasma samples from fasting EDTA-treated blood was frozen immediately upon collection at −80°C until aliquoting. Upon thawing, aliquoted samples were completely randomized prior to processing for mass spectrometry analysis. 17 steroid hormones were measured using the AbsoluteIDQ Stero17 kit (Biocrates, Austria), which included glucocorticoids (cortisol, cortisone, and 11-deoxycortisol), mineralocorticoids (aldosterone, corticosterone, and deoxycorticosterone), progestogens (17α-hydroxyprogesterone and progesterone), estrogens (estrone and estradiol), and androgens (androstenedione, androsterone, dehydroepiandrosterone, dehydroepiandrosterone sulfate, dihydrotestosterone, etiocholanolone, and testosterone). Nanomolar concentrations were obtained from Biocrates in an Excel sheet along with a corresponding data report. Given that the measurements were taken across multiple plates, Biocrates calculated a project-based limit of detection (LOD), corresponding with the highest LOD in any plate for each steroid hormone. For measurements below LOD, Biocrates imputed missing values using the R package *logspline*. The *logspline* imputation method generates values between LOD and LOD/2 and applies the variance between values above LOD. Therefore, if there is high variance in the dataset, the imputed values may also exceed these limits. In this study, the steroid hormones were analyzed together with the electronic case report form factors for their influence on body temperature.

##### Peripheral immune cell parameters

Ten eight-color immunophenotyping flow-cytometry panels were used on unstimulated blood to report a total of 166 distinct immunophenotypes, including counts, ratios, and mean fluorescent intensities of relevant markers as previously reported^30^.

##### Milieu Interieur *ex-vivo* stimulations and subsequent transcriptomic and proteomic assays

TruCulture stimulations were performed as previously described^31^. Briefly, TruCulture tubes were prepared in batch for each stimulus, resuspended in a volume of 2 mL buffered media, and stored at −20°C until time of use. On day of sampling, 1 mL of whole blood was distributed into 37°C prewarmed TruCulture tubes for each stimulus within 15 minutes then incubated for 22 hours at 37°C in a dry block. Post-incubation, tubes were opened and a valve was inserted to separate the cells from the supernatant and stop the stimulation. Supernatants were aliquoted and frozen at −80°C until assayed and RNA samples were processed as previously described^32^. Transcriptomic profiling of the immune cells was completed as previously described using the NanoString Human Immunology v2 Gene Expression CodeSet^4^. Proteomic profiling of the supernatants was completed by Rules Based Medicine (Austin, Texas, USA) using Luminex xMAP technology^33^. This paper analyzed four stimulation conditions: bacterial (E. coli; TLR 2, 4 agonist), viral (Poly I:C, TLR3, RIG-I, MDA-5 agonist), superantigen (SEB, T cell agonist), and a nonstimulated control.

##### COVIMUN cohort *ex vivo* stimulations and subsequent transcriptomic and proteomic assays

Assessment of 82 patients from a Danish cohort of COVID-19 patients included gene expression profiling in stimulated and unstimulated whole blood. At hospital admission during the early pandemic, 1 mL of whole blood collected in lithium heparin tubes was transferred to prewarmed TruCulture tubes (Q2 Solutions, USA) containing no stimulus (negative control) as previously described^61^. Following 22h incubation at 37 °C, supernatants were separated after addition of 2 mL TRIzol™ Reagent (Thermo Fisher Scientific, USA) for RNA preservation, and cell pellets were stored at −80 °C until RNA extraction. Assessment of RNA extraction from TruCulture blood tubes was performed using a custom pre-treatment protocol followed by the NucleoSpin 96 RNA Kit (Machery-Nagel, Germany). Samples were thawed on ice for 60 min, vortexed (2 × 5 min, 2000 rpm), and centrifuged (3000 × g, 5 min, 4 °C). RNA was eluted in 50 µL RNase-free H₂O, quality controlled using an Agilent TapeStation (Agilent, USA) and normalized to 7 ng/µL.

RNA extracted from unstimulated samples from 102 patients were analyzed using NanoString nCounter Human Immunology v2 panels (NanoString Technologies, Inc., USA), targeting 579 genes involved in innate and adaptive immune pathways and 15 housekeeping genes. A total of 56 ng RNA per sample was hybridized according to the MAN-10056-03 (January 2020) protocol, with a modified input volume of 8 µL RNA. Samples were incubated with reporter and capture probes for 24 h and analyzed on the NanoString nCounter Flex system. All cartridges were scanned at an FOV count of 555. Preprocessing and quality control of NanoString nCounter raw data were performed using a previously described approach^62^ implemented with the *RUVSeq3*^63^ R package, with housekeeping genes selected using the *geNorm* function from the *ctrlGene* package. Following QC and normalization, 14 patients were excluded due to either flagging by nCounter or *RUVSeq* flagging or data missingness. Of the 88 remaining patients, 82 had temperature data noted in their electronic health record data and were included for downstream analyses.

Clinical patient data included classification of disease severity: Grade 1, <3 L O₂ to maintain SpO₂ >92%; Grade 2, >3 L O₂ to maintain SpO₂ >92%; Grade 3, >6 L O₂ to maintain SpO₂ >92% and/or ICU admission; and Grade 4, mechanical ventilation and/or death. Immunosuppressive status encompassed clinical conditions and medications known to impair immune function.

#### Data generation and analysis

Unless otherwise stated, all displayed MI results were performed on the 949 individuals of the cohort who gave consent to share their data publicly. All regression model effect size plots are on a linear scale and display the extracted estimate and 95% confidence interval. Effects with significant FDR-corrected q values (<0.05) are in black, and those that did not reach significance are in grey.

##### Temperature Variation analysis

Multiple regression models were built to estimate the effects of intrinsic and behavioral factors on body temperature. In all donors, the model included the covariates of age, sex, and their interaction. In pre-menopausal donors, the covariates of age and oral contraceptive use were included, and for females between 44-55 years, age and menopause were included. A multiple regression model with dispersion formula was built to estimate the effects of age, sex, their interaction, periodic time of day, and periodic day in the year on the variance in body temperature over 3 visits spanning 2-8 weeks and applied using the R *glmmTMB* package.

##### Electronic case report form factor analysis

To test for associations between demographic variables and steroid hormone levels we used the R package *stats* to perform likelihood ratio tests comparing a base model controlling for age, sex and an age×sex interaction [lm(temperature ∼ age×sex)], with a model controlling for age, sex and an age sex interaction plus a predictor demographic variable [lm(temperature ∼ age×sex + demographic variable)]. We performed this analysis on 137 demographic variables in all donors, then for male and female donors independently. The independent sex models still controlled for age, with 14 additional variables in the female models related to contraception, hormonal treatment, and family history of reproductive cancer. An FDR correction was performed to account for multiple testing across all demographic features for each hormone. Heatmaps to display the q value from the LRT were generated using the R package *pheatmap*.

##### Peripheral immune cell composition analysis

To describe natural variation of immune cell counts, ratios, and mean fluorescent intensities (MFIs) of relevant cell surface proteins, unstimulated blood was stained with ten unique immunophenotyping flow-cytometry panels were used on unstimulated blood to report a total of 166 distinct immunophenotypes^30^. We applied linear mixed models to these parameters with body temperature as a covariate while controlling for sex, age, age×sex interaction, smoking, body mass index, time of sampling, and day of sampling as a random effect, as well as CMV status when modeling cell counts. Similar models were applied to all donor data, with additional covariates for body temperature and the age×sex interaction, as well as a spline fitted to time of sampling to account for potential non-linear effects. These covariates were selected as previously reported, with the addition of the age-sex interaction, body temperature, and a spline for time of sampling^30^. Multiple testing correction was performed using for FDR (FDR q < 0.05 considered significant) and models were applied using the *MIIC* R package^30^.

#### *Ex vivo* immune response analysis

To model temperature effects on *ex-vivo* stimulation responses (bacterial (E. coli; TLR 2, 4 agonist), viral (Poly I:C, TLR3, RIG-I, MDA-5 agonist), superantigen (SEB, T cell agonist), nonstimulated control), linear models were applied to induced cytokine and gene expression data (log2FC over the nonstimulated control). Covariate selection: all models included age, periodic time and day of sampling and 16 immune cell proportions including neutrophils, basophils, eosinophils, monocytes, B cells, NK cells, dendritic cells (DCs), and 9 T cell subsets of different naïve, effector and memory CD4 and CD8 cells. Smoking and CMV were added as necessary whether they contributed to the models as determined by LRT. For *E. coli* and Poly I:C, smoking was significant for some cytokines by drop1(model, test = "Chi"), but CMV was not. Both were sometimes significant for SEB cytokines, so they were included in the models for SEB. Multiple testing correction was performed within each sub-cohort using FDR (FDR q < 0.05 considered significant).

##### Healthy Donor Blood and Stimulation

Human peripheral blood samples were collected from healthy volunteers through the Clinical Investigation Center INVOLvE (Investigation and volunteers for human health) of the Institute Pasteur under project M22 and COSIPOP 2023-30 (NCT03925272) through the Établissement français du sang (EFS). The participants received oral and written information about the research and gave written informed consent in the frame of the CoSImmGEn cohort after approval of the CPP Ile-de-France I Ethics Committee (2011, jan 18th) and the COSIPOP cohort after approval of the CPP Est II Ethics Committee (2023, feb 20th). Blood was collected in heparin tubes and delivered within 6 hours. In round-bottom 96-well plates, 100 μL of blood was added to 200 μL of pre-warmed TruCulture media for each condition in duplicate or triplicate (bacterial: LPS component of E. coli; TLR4 agonist; viral: R848 (TLR7/8 agonist); superantigen (CytoStim, T cell agonist); and a nonstimulated control). Plates were placed in cell incubators set to 0% CO_2_, 0.02% O_2_, and 36.8°C or 39°C respectively for 22 hours resulting in 8 experimental conditions per donor (2 temperatures × 4 stimulations). Post-incubation, plates were centrifuged at 300g for 5 minutes. The supernatant was collected and stored at −20°C until time of use. Cells were either assayed by flow cytometry to confirm viability using a Fixable Viability Dye eBioscience eFluor 506, or diluted 2:1 with Trizol, shaken for 2 minutes at 2000 rpm, rested for 10 minutes at room temperature, then frozen at −80°C until time of use.

For the single-cell experiments, 1 mL of blood was added to prewarmed TruCulture tubes for each stimulus (superantigen (CytoStim; T cell agonist), viral (Poly I:C; TLR3, RIG-I, MDA-5 agonist), bacterial (*E. coli*; TLR2/4 agonist), nonstimulated control) and incubated at 36.8°C-37°C or 39°C respectively for 4 hours in dry blocks. Post-incubation, tubes were opened and a valve was inserted to separate the cells from 1.5 mL of supernatant and stop the stimulation. Supernatants were aliquoted and frozen at −80°C until assayed and the remainder of the 1.5 mL volume was layered onto 2 mL Ficoll in 5 mL tubes and spun for 20 minutes at 800g without braking to separate the PBMCs. We proceeded to wash the PBMCs twice in 15 mL conical tubes with 10 mL of PBS and spins of 5 minutes at 400g and resuspended the pellet in 1 mL PBS + 0.04% BSA for cell counting. The cells from each pool were counted using the Cell Countess 3 automated cell counter (Thermo Fisher Scientific) and the cell density was adjusted to 1,000 viable cells per µl of PBS + 0.04% BSA and we proceeded with 10X demonstrated protocols described below.

##### LEGENDplex Cytokine assay and analysis

On day of assaying, supernatants were thawed and centrifuged at 300g for 5 minutes. The concentration of 14 different analytes was measured using the LEGENDplex Human Covid-19 Cytokine Storm Panel 1(14-plex) kits and the manufacturer protocol was followed (BioLegend, San Diego, California, USA). Plates were read on a BD Fortessa II 5-color flow cytometer (Franklin Lakes, New Jersey, USA), and results were analyzed on the BioLegend portal (https://legendplex.qognit.com/). Concentrations were log_2_-transformed and controlled for donor and LEGENDplex experiment batch effects via linear modeling with the package *lme4*. Median controlled concentrations were found for each analyte per stimulation condition, and the log_2_FC of the stimulation over the nonstimulated control was calculated. Analytes with a log_2_FC of >1 were considered induced and were further analyzed for temperature effects. To achieve this, linear mixed models were applied to log_2_-transformed concentrations using the *lme4* package with the covariates of stimulation, temperature, a stimulation×temperature interaction, and the random effects of donor and LEGENDplex experiment. Multiple testing correction was performed within each stimulation using FDR (FDR q < 0.05 considered significant). To ensure the observed temperature-induced decreases in cytokine levels in the viral and bacterial conditions were not due to cell death or protein instability, we isolated cells at 22 hours and performed flow cytometry with a fixable viability dye and bulk RNA sequencing (RNAseq). Flow cytometry confirmed comparable cell viability across all stimuli in both temperature conditions with averages of 89.8% at 36.8°C and 89.9% at 39°C (**Supp. Fig. 3B**).

##### RNA sequencing and analysis

On day of extraction, cells in Trizol were thawed, and 2 100-μL aliquots per condition per donor were combined for extraction. RNA extraction was performed using the PureLink RNA Mini Kit (ThermoFisher Scientific, Waltham, Massachusetts, USA) per the manufacturer protocol with the optional DNase step and quantified with the Qubit High Sensitivity RNA assay (ThermoFisher Scientific, Waltham, Massachusetts, USA) and qualified with the Agilent High Sensitivity RNA ScreenTape (Agilent Technologies, Santa Clara, California, USA). RNA was sequenced via the BRBseq protocol by Alithea Genomics (Épalinges, Switzerland). The analysis was modeled after published methods using the *edgeR*, *limma*, *glimma*, and *camera* packages^64^. Using the *edgeR* package, initial filtering and normalization of the UMI-corrected counts per gene was completed and the log_2_ fold change of the LPS condition over the nonstimulated control condition was calculated. Using the R *limma* package, differentially expressed genes between the 36.8°C and 39°C condition were calculated using empirical Bayes moderated t-statistics. We then applied the *camera* method to the significant differently expressed genes for gene set testing with the Broad Institute’s MSigDB hallmark gene sets^65^ and multiple testing correction was performed within each stimulation using FDR (FDR q < 0.05 considered significant).

##### Single-cell RNA and cell surface marker sequencing

Isolated PBMCs were labeled for CITE-seq according to the manufacturer’s instructions demonstrated protocol CG000149 (10x Genomics). Washed PBMCs were resuspended in chilled 1% BSA in phosphate-buffered saline and incubated with human TruStain FcX blocking solution (BioLegend) for 10 min at 4 °C. Cells were then stained with a cocktail of 20 TotalSeq-B antibodies (BioLegend) previously centrifuged at 14,000*g* for 10 min (**Supplementary Table 2**). The cells were incubated for 30 min at 4 °C in the dark and then washed three times. Cell concentration was again determined using Cell Countess 3 automated cell counter (Thermo Fisher Scientific) and density was then adjusted to 1,000 viable cells per µl in 1% BSA in phosphate-buffered saline. We generated scRNA-seq libraries and cell protein libraries with the Chromium Single Cell 3′ Reagent Kit (v.4), using the Feature Barcoding technology for Cell Surface Proteins (10x Genomics).

##### Single-cell RNA sequencing data processing and quality control

Single-cell RNA sequencing data from peripheral blood mononuclear cells (PBMCs) were processed using *scanpy* (v1.9.3) in Python 3.11. Raw count matrices were loaded and merged across all samples, retaining cells with unique molecular identifier (UMI) counts between 200 and 30,000, and genes expressed in at least 3 cells. Doublets were identified and removed via *scrublet*. Initial filtering removed cells with mitochondrial gene content exceeding 15%, hemoglobin content exceeding 10%, and less than 300 genes. After clustering and cell type annotation, a second round of filtering removed cells with mitochondrial gene content exceeding the 95^th^ percentile (5.3%) and less than the 5^th^ percentile of number of genes (712). After quality filtering, the dataset comprised of 640,871 cells and 3,396 genes from 4 healthy donors.

##### Cell type annotation

Cell types were assigned using a reference-based approach with pre-trained models from Cell Typist (https://www.celltypist.org/)^66^. The final model applied was trained on 1.5 million stimulated PBMCs. Initial annotations were refined by examining canonical marker gene expression patterns including CD3 (T cells), CD20 (B cells), CD14/CD16 (monocytes), CD56 (NK cells), and lineage-specific markers. Rare cell populations including innate lymphoid cells (ILCs), plasmacytoid dendritic cells (pDCs), and AXL+ SIGLEC6+ dendritic cells (ASDCs) were identified based on specific marker combinations. Cell type assignments were validated through UMAP visualization, as well as marker gene and cell surface protein expression analysis.

##### Pseudobulk differential gene expression and gene set analysis

To leverage the statistical power of bulk RNA-seq methods while accounting for biological replication, we performed pseudobulk analysis by aggregating gene counts within each cell type, donor, and experimental condition combination (stimulation and temperature). Only cell type-condition combinations with at least 5 cells were retained for analysis. Gene expression was normalized using DESeq2’s median of ratios method, and genes with mean expression <1 TPM across conditions were filtered out.

Differential expression analysis was performed using DESeq2 (v1.38.0) with the design formula: ∼ donor + condition. Comparisons were made between temperatures within each stimulation condition. Genes were considered significantly differentially expressed at FDR-adjusted p-value < 0.05 and |log2 fold change| > 0.2. We then applied the *camera* method to the significant genes for gene set testing with the Broad Institute’s MSigDB hallmark gene sets^65^. Pathways with fewer than 5 genes represented in the expression matrix were excluded from testing and analysis was completed for all donors and on a sex-stratified basis. Gene set test results were corrected for multiple testing, with FDR < 0.05 considered statistically significant. To quantify biological magnitude beyond statistical significance, we calculated a "collective beta" (Pathway LogFC) for each pathway. This metric represents the mean gene-level log₂ fold-change coefficient across all genes within a Hallmark gene set: β_(pathway) = 1/|S| ∑_(g∈S) β_g. This provides an interpretable effect size metric where a pathway beta of 1 indicates an average 2-fold upregulation of pathway gene. To identify cell types exhibiting the greatest sex differences in pathway responses, we computed a Sex Divergence Score for each cell type across superantigen, viral, and bacterial stimulation conditions. For each pathway, we calculated: Divergence_(pathway) = |β_(female) −β_(male)| × −log_10 (min(FDR_(female), FDR_(male))). Cell types were ranked by the sum of divergence scores across all Hallmark pathways and stimulation conditions. The top 10 cell types were selected for focused visualization, ensuring representation of both statistically confident and biologically substantial sex differences.

##### COVIMUN cohort gene expression analysis

Raw NanoString gene expression counts were processed using the *edgeR* and *limma* packages in R. Genes with low expression were filtered using the *filterByExpr* function to retain only genes with sufficient counts per million (CPM) in a minimum number of samples. Library size normalization was performed using the Trimmed Mean of M-values (TMM) method to correct for compositional biases. We used a linear modeling approach with *limma-voom* to identify genes associated with clinical temperature measurements. The normalized counts were transformed using *voomWithQualityWeights* to estimate the mean-variance relationship and assign observational quality weights. A design matrix was constructed adjusting for potential confounders, including age, disease severity, sex, and time since symptom onset (modeled using sine and cosine transformations to account for periodicity).

Body temperature was analyzed as a continuous variable (°C) or as a binary fever status (febrile: > 37.5°C; non-febrile: ≤ 37.5°C) in the total cohort and in a sex-stratified manner. Linear models were fitted using *lmFit*, and empirical Bayes moderation was applied using *eBayes* to stabilize variance estimates. P-values were adjusted for multiple testing using the Benjamini–Hochberg (BH) method to control the False Discovery Rate (FDR). To visualize the transcriptional signature associated with fever, a subset of the top differentially expressed genes (DEGs) was selected based on the lowest FDR-adjusted p-values. For the male-specific heatmap, expression values were row-scaled to Z-scores to standardize gene expression across samples. Hierarchical clustering of genes (rows) was performed using Euclidean distance and complete linkage. Samples (columns) were ordered via supervised clustering based on fever status to facilitate the visual comparison of febrile versus non-febrile profiles. Heatmaps were generated using the *pheatmap* package, with clinical annotations (severity and fever) displayed alongside the expression matrix.

##### ReCoN multicellular network analysis

ReCoN models multicellular systems by assembling a heterogeneous multilayer network that encodes molecular interactions across cell types^67^. It comprises a gene regulatory network (GRN) between TF and genes, a receptor-to-gene network, and the cell-cell communication network between produced ligands and receptors. Signals can be propagated through the network using random walks with restarts. The final score assigned to each node reflects regulatory influence, or as estimated by the stationary distribution of the random walk seeded from genes of interest in the receiver cell type.

From the gene expression of some cell types (called sender cells) at 4h, ReCoN was used to predict the future gene expression of cell types (called receiver cells) in male when exposed to Poly I:C stimulation and to recover the molecular cascades explaining these transcriptomic profiles. For both temperature conditions, febrile (39°C) and healthy (36.8°C), the same network, seeds, and parameters were used. We prioritized male-biased, temperature-sensitive cell populations and genes, defined by differential expression exceeding both negative control and female viral responses (see **ReCoN selection of sender and receiver cell types and seed genes** below). The weight attributed to each seed was the average expression of the corresponding genes in the corresponding cell type in the corresponding condition. Two explorations were run in each condition, to retrieve the upstream cascades in the sending cells and the downstream cascades, through cell communication, in the receiving cell type.

All analyses were run with the default parameters of ReCoN: a restart probability of 0.6, equiprobable transitions between network layers, and the default exploration config “intercell_upstream”. ReCoN analyses were conducted for each combination of sex, temperature, and immune stimulus, and submitted as parallel jobs on a SLURM computing cluster.

##### ReCoN network construction

A common gene regulatory network was generated from all cells across conditions with the GRNBoost2 (implemented in *arboreto* v0.1.6)^68^, and the receptor-to-gene network called human_receptor_gene_from_NichenetPKN from ReCoN was used for all analyses. A cell-cell communication (CCC) network was inferred independently for each combination of sex, temperature, and immune stimulus using the *LIANA+* package^69^, applying the *CellPhoneDB*^70^ algorithm with the *human_consensus* ligand-receptor database. Cell types lacking detectable ligand or receptor expression under a given condition were excluded from the corresponding network. To derive a stimulus-level consensus, CCC scores were averaged across temperature conditions within each stimulus and visualized as directed chord diagrams, in which arc width encodes the total aggregated communication score between each pair of cell types. Chord diagrams were generated with *Circlize* ^68^.

##### ReCoN selection of sender and receiver cell types and seed genes

To identify candidate ligands and receptors for cell-cell communication (CCC) analysis, we implemented a multi-step gene selection pipeline using sex-stratified single-cell pseudobulk differential expression results. Our objective was to isolate genes whose temperature responsiveness was specific to male viral immune responses. First, we identified temperature-responsive genes specific to the stimulation condition and sex using a Difference-in-Differences (DiD) approach. For each gene, we calculated a specificity score defined as the difference between the temperature log2 fold-change in the male Poly I:C condition and the temperature LFC in the male Negative Control condition, as well as the temperature log2 fold-change in the male and female Poly I:C condition. Genes were retained if they were significantly differentially expressed (FDR <0.05) with temperature in the Poly I:C condition, had a 15% fold-change difference than the male negative control condition (DiD score > 0.2), and had a sex-specific response (FDR > 0.10 in the female Poly I:C temperature contrast or DiD score > 0.2). This pipeline was applied independently across all 42 annotated cell types. To ensure sufficient signal for downstream CCC inference, cell types yielding fewer than 3 selected genes were excluded from further analysis. We prioritized a maximum of 20 cell types for ReCoN analysis, and selected sender and receiver cell types for the male viral condition through their known role in the literature, cell type abundance, and their quantitative importance in the CCC network. All analyses were performed using Python with the *pandas* and *numpy* packages.

##### ReCoN contrast analysis and cascade visualization

Log Fold Changes were computed for the normalized ReCoN scores from febrile (39°C) and healthy (36.8°C-37°C) explorations to identify differences across both conditions. Gene sets corresponding to the selected pathways were retrieved from the MSigDB Hallmark collection (v2020)^72^ using *camera*^73^. The full signaling cascade was then shown for the ligands with the most sex-specific temperature sensitivity. Each ellipse in the diagram corresponds to a distinct molecular interface. The node colors are defined by whether expression increased with lower temperature (36.8°C-37°C, blue) or higher temperature (39°C, red). Regulatory links follow the color of the upstream node. For visualization purposes, we selected in the figure the three receiver cells most biologically relevant to viral immune response with the greatest sex differences in pathway signaling. Sender cells were identified as those contributing the largest fraction of ligand signal directed toward the selected receivers. To characterize intercellular cascades in full, an extended diagram additionally incorporates the upstream regulatory context of the sender cell, tracing mechanisms from sender-cell receptors through transcription factors to the secreted ligands.

#### Data and materials availability

All published Milieu Intérieur pseudonymized datasets can be accessed on https://dataset.owey.io/ by submitting a data access request at https://redcap.pasteur.fr/surveys/?s=ND8TP8MDD3. The request will be reviewed by the Milieu Intérieur data access committee (DAC). The DAC informs the research participants of the data access request and grants data access if the request is consistent with the informed consent signed by the participants. Research on Milieu Intérieur datasets is restricted to research on the genetic and environmental determinants of human variation in immune responses. Data access is typically granted two months after request submission. Experimental data sets will be made available upon publication.

## Data Availability

All data produced in the present study are available upon reasonable request to the authors

https://dataset.owey.io/

https://redcap.pasteur.fr/surveys/?s=ND8TP8MDD3

## Acknowledgements

Elizabeth Maloney was supported by a stipend from the Pasteur - Paris University (PPU) International PhD program and by the LabEx *Milieu Intérieur* (French government’s Invest in the Future programme; reference ANR-10-LABX-69-01) and is a student from the FIRE Ph.D. program funded by the Bettencourt Schueller Foundation and the EURIP Graduate Program (ANR-17-EURE-0012), and received doctoral support from INCEPTION. Rémi Trimbour was supported by the French government’s Agence Nationale de la Recherche under the “France 2030” program (ANR-23-IACL-0008, PRAIRIE-PSAI). The COVIMUN study was funded by a COVID-19 grant from the Independent Research Fund Denmark, Ministry of Higher Education and Science (0238-00006B). We are grateful to the CRBIP unit CHIP for providing fit-for-purpose biological resources and associated services and to the healthy volunteers of the CoSImmGEn and COSIPOP cohorts who agreed to the scientific use of their samples and data. We thank the Institut Pasteur Single Cell Biomarker and Sequencing platforms for access to technologies used in this study. We also thank Aurélie Bisiaux for CITE-seq protocol sharing and Jean-Marc Doisne for Fortessa sharing. We thank Emeline Perthame, Vincent Rouilly, Jacob Bergstedt, and Rafael De Andrade Moral for modeling advice, and Auxence Desrentes, Yann Aquino and Gaston Rijo de Leon for code sharing. Thanks to Anthony Bertrand for R and RNAseq analysis support, and Stella Hoft and Jessica Mathews for editing support. Thanks also to Louise Dupuis and Herve Isambert for causal analysis discussions. Special thanks to Molly Ingersoll and her lab for their enthusiastic support. Another special thank you to Vincent Bondet for lab equipment management and our entire lab for support and advice throughout the project.

## Contributions

E.M. conceived the study, performed analysis and experiments, wrote and edited the manuscript. R.T. performed the ReCoN analysis underlying Figure 7 and generated parts A and B. M.S. supported single-cell analysis with consultations, code, methods and visualization advice. J.S. performed genetic analysis which was not included in the final manuscript and provided data interpretation feedback throughout the project. P.T.B. performed COVIMUN gene expression analysis underlying Figure 6, generated part E, and earlier versions of parts B-D. E.V. used a novel algorithm to identify bridge samples across the bulk proteomic assay plates in the *ex vivo* fever experiments. The COSIPOP study group provided healthy donor samples for the *ex vivo* fever experiments. R.S.T., M.E.E.M., B.L., A.O.G., S.D.N., D.P., C.U.N., S.R.O. and the COVIMUN study consortium conducted the COVID-19 clinical study analyzed in Figure 6. M.R. provided advice on linear regression and mixed modeling. E.P. provided advice on hormone mediation analysis. L.Q.M. obtained funding and coordinated the clinical samples. D.D conceived and supervised the study, edited the manuscript, obtained funding and coordinated the clinical samples.

The Milieu Intérieur Consortium¶ is composed of the following team leaders: Laurent Abel (Hôpital Necker), Andres Alcover, Hugues Aschard, Philippe Bousso, Nollaig Bourke (Trinity College Dublin), Petter Brodin (Karolinska Institutet), Pierre Bruhns, Nadine Cerf-Bensussan (INSERM UMR 1163 – Institut Imagine), Ana Cumano, Christophe D’Enfert, Caroline Demangel, Ludovic Deriano, Marie-Agnès Dillies, James Di Santo, Gérard Eberl, Jost Enninga, Jacques Fellay (EPFL, Lausanne), Ivo Gomperts-Boneca, Milena Hasan, Gunilla Karlsson Hedestam (Karolinska Institutet), Serge Hercberg (Université Paris 13), Molly A Ingersoll (Institut Cochin and Institut Pasteur), Olivier Lantz (Institut Curie), Rose Anne Kenny (Trinity College Dublin), Mickaël Ménager (INSERM UMR 1163 – Institut Imagine), Frédérique Michel, Hugo Mouquet, Cliona O’Farrelly (Trinity College Dublin), Etienne Patin, Antonio Rausell (INSERM UMR 1163 – Institut Imagine), Frédéric Rieux-Laucat (INSERM UMR 1163 – Institut Imagine), Lars Rogge, Magnus Fontes (Institut Roche), Anavaj Sakuntabhai, Olivier Schwartz, Benno Schwikowski, Spencer Shorte, Frédéric Tangy, Antoine Toubert (Hôpital Saint-Louis), Mathilde Touvier (Université Paris 13), Marie-Noëlle Ungeheuer, Christophe Zimmer, Matthew L. Albert (Octant Biosciences), Darragh Duffy§, Lluis Quintana-Murci§,

¶ unless otherwise indicated, partners are located at Institut Pasteur, Paris

§ co-coordinators of the Milieu Intérieur Consortium

Additional information can be found at: http://www.milieuinterieur.fr

The COSIPOP study group is constituted by members who designed and manage the COSIPOP cohort from the Institut Pasteur with the following persons: AROWAS Laurence, CLEMENT Nathalie, DELHAYE Maurine, FANAUD Christine, NGUYEN Emilie, NOURY Clémence, PERLAZA Blanca-Liliana, VOGTENSPERGER Marie, ZAYOUD Ayla and lastly JOLLY Nathalie and LAUDE Hélène, as co-senior authors *\**

The COVIMUN gene expression cohort study group is composed of the following team members with affiliations at the Copenhagen University Hospital and/or the department of Clinical Medicine or Department of Clinical Biochemistry at the University of Copenhagen: Tereza Faitova, Adin Sejdic, Zitta Barrella Harboe, Ditte Stampe Hersby, Christian Brieghel, Annemette Hald, Hanne Marquart, Jens Lundgren, Anne-Mette Lebech, Marie Helleberg, Frederik Otzen Bagger.

## Supplementary Information

### List of Supplementary data

**Supplementary Figure 1.**
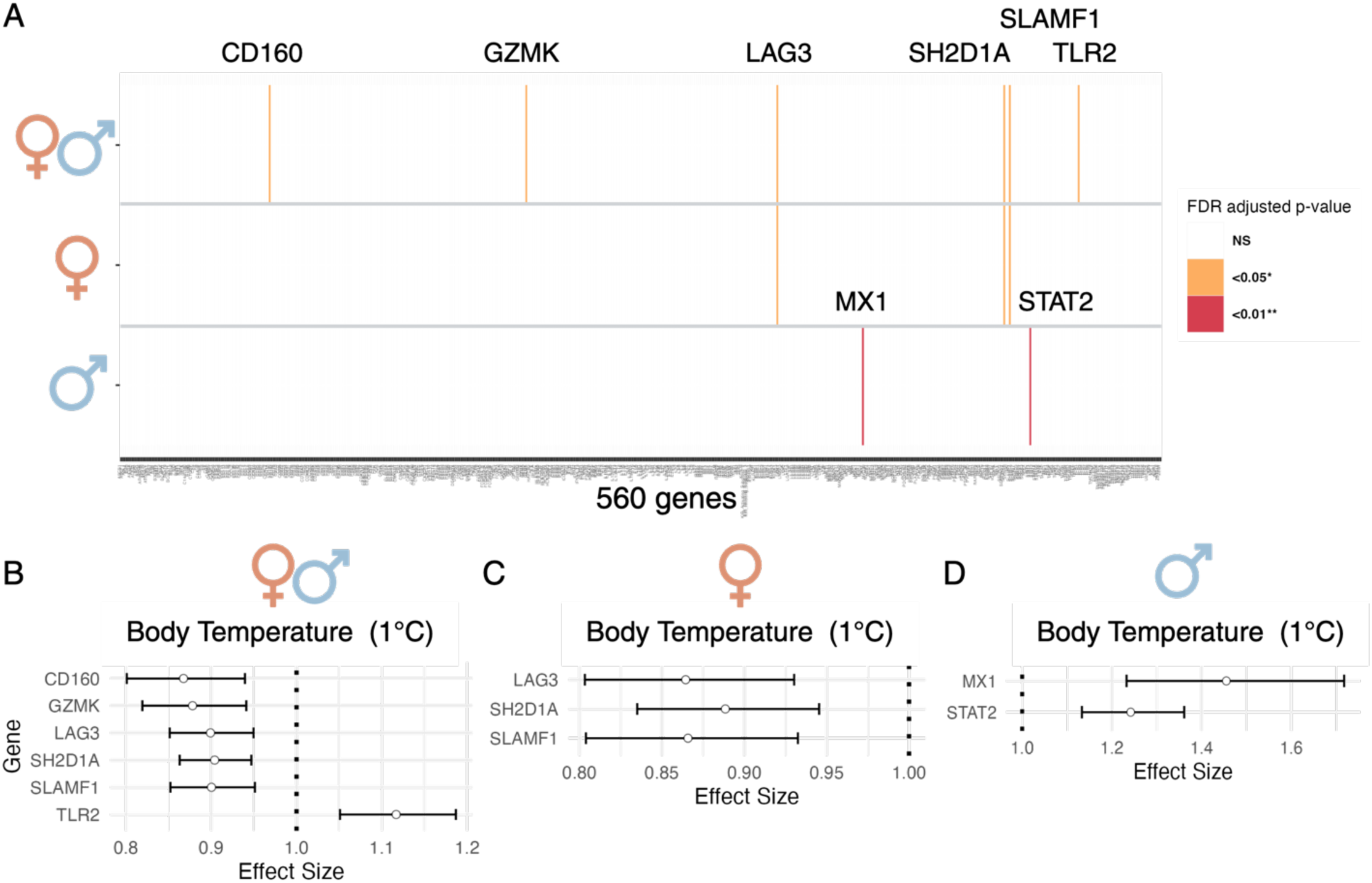
Body temperature effects on negative control condition immune responses. **A** Barcode plot of genes whose expression was significantly associated with body temperature from multiple regression models applied to NanoString gene expression data from the nonstimulated condition in all, female, or male donors. Models controlled for age, 16 major cell subtype proportions, periodic time and day of sampling, with the addition of sex and an age×sex interaction for all donors. FDR *q*<0.05 in orange, *q*<0.01 in red. Quantification of the effect of body temperature on gene expression estimated from these models, then transformed to the original scale in **B** all, **C** female, and **D** male donors. (n=949 (all donors), n=477 (males), n=472 (females))

**Supplementary Figure 2.**
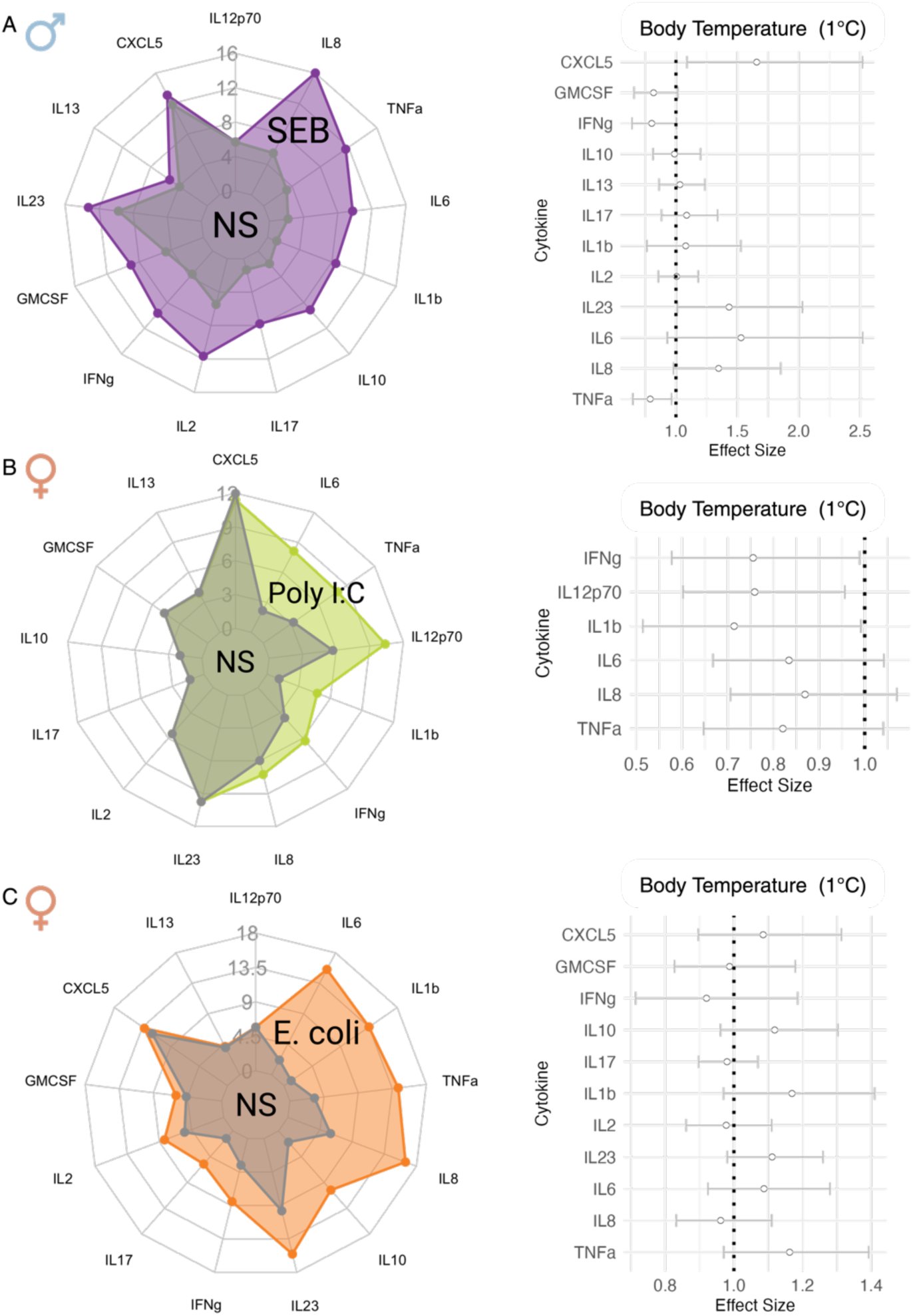
No significant body temperature effects on adaptive immune responses in male donors and innate immune responses in female donors. Spider plot showing cytokine levels in nonstimulated (grey) and **A** SEB (purple), **B** Poly I:C (green) or **C** *E. coli* (orange) condition; cytokines were considered induced if the levels in the stimulation condition were doubled from the nonstimulated control. **Right** Quantification of the effect of body temperature on log2-transformed induced cytokine levels, estimated in a multiple regression model controlling for age, 16 major cell subtype proportions, periodic time and day of sampling, smoking status, and CMV for the SEB condition, then transformed to the original scale. Effect sizes with FDR corrected *q*-values < 0.05 are in black, nonsignificant effects in grey.

**Supplementary Figure 3.**
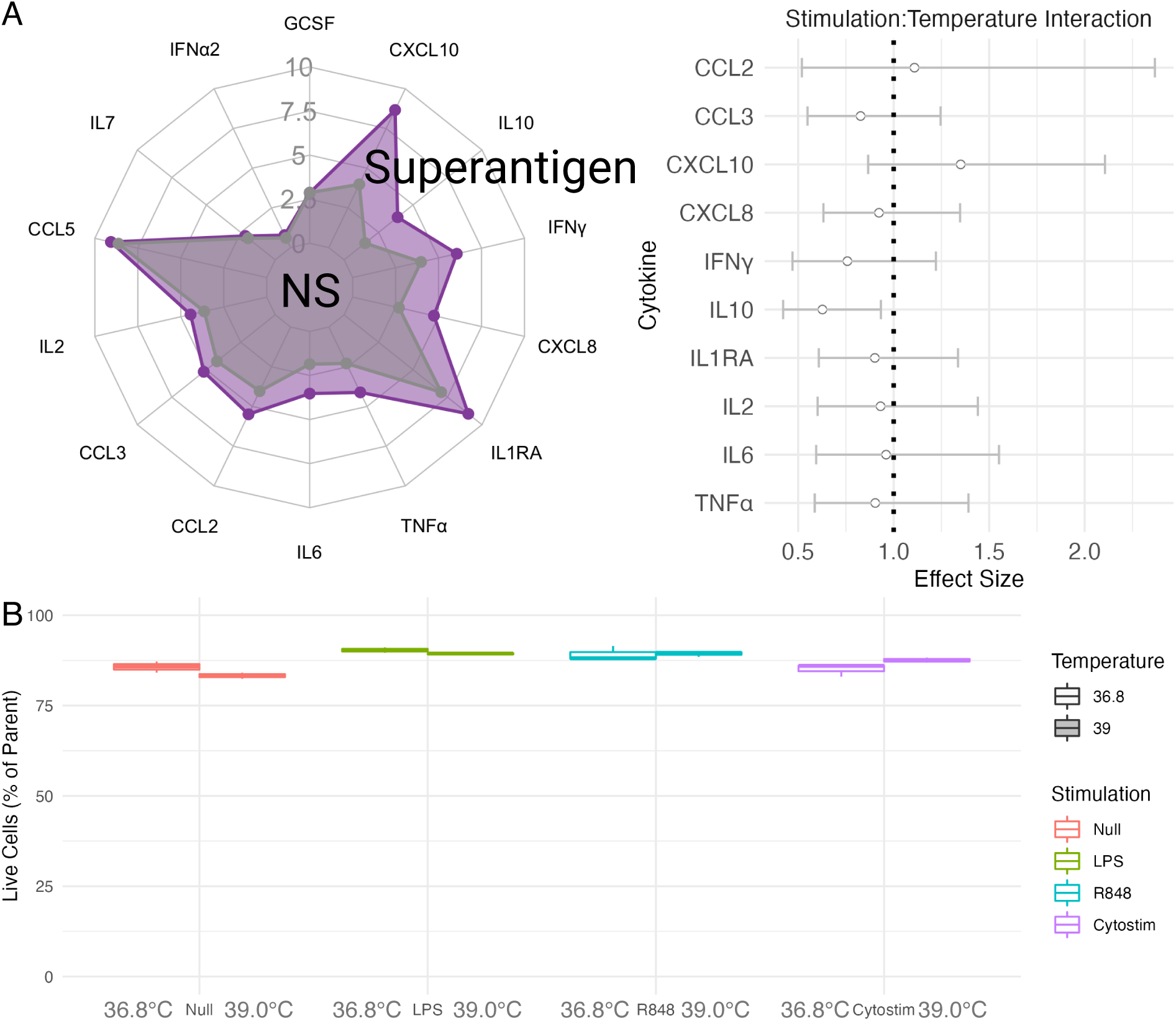
Febrile temperature does not significantly influence cytokine levels in CytoStim *ex vivo* whole blood stimulation or cell viability. **A** Spider plot showing median cytokine levels in the supernatant of healthy whole blood stimulations (n = 20) after 22 hours of incubation in the nonstimulated (grey) or CytoStim (purple) condition; cytokines were considered induced if the median levels in the stimulation condition were doubled from the nonstimulated control. **Right** Quantification of the interaction effect of stimulation and incubation temperature (36.8°C or 39°C), on the abundance of induced cytokines estimated in a linear mixed model with log2-transformed concentration as the response, controlled for donor and batch effect, then transformed to the original scale. Effect sizes with FDR corrected q-values < 0.05 are in black, nonsignificant effects in grey. **B** Boxplot showing percent live whole blood cells after 22 hours of incubation at 36.8°C (left) or 39°C (right), in all four stimulations (nonstimulated null control in red, LPS in green, R848 in blue, CytoStim in purple). Assayed using flow cytometry and a Fixable Viability Dye.

**Supplementary Figure 4.**
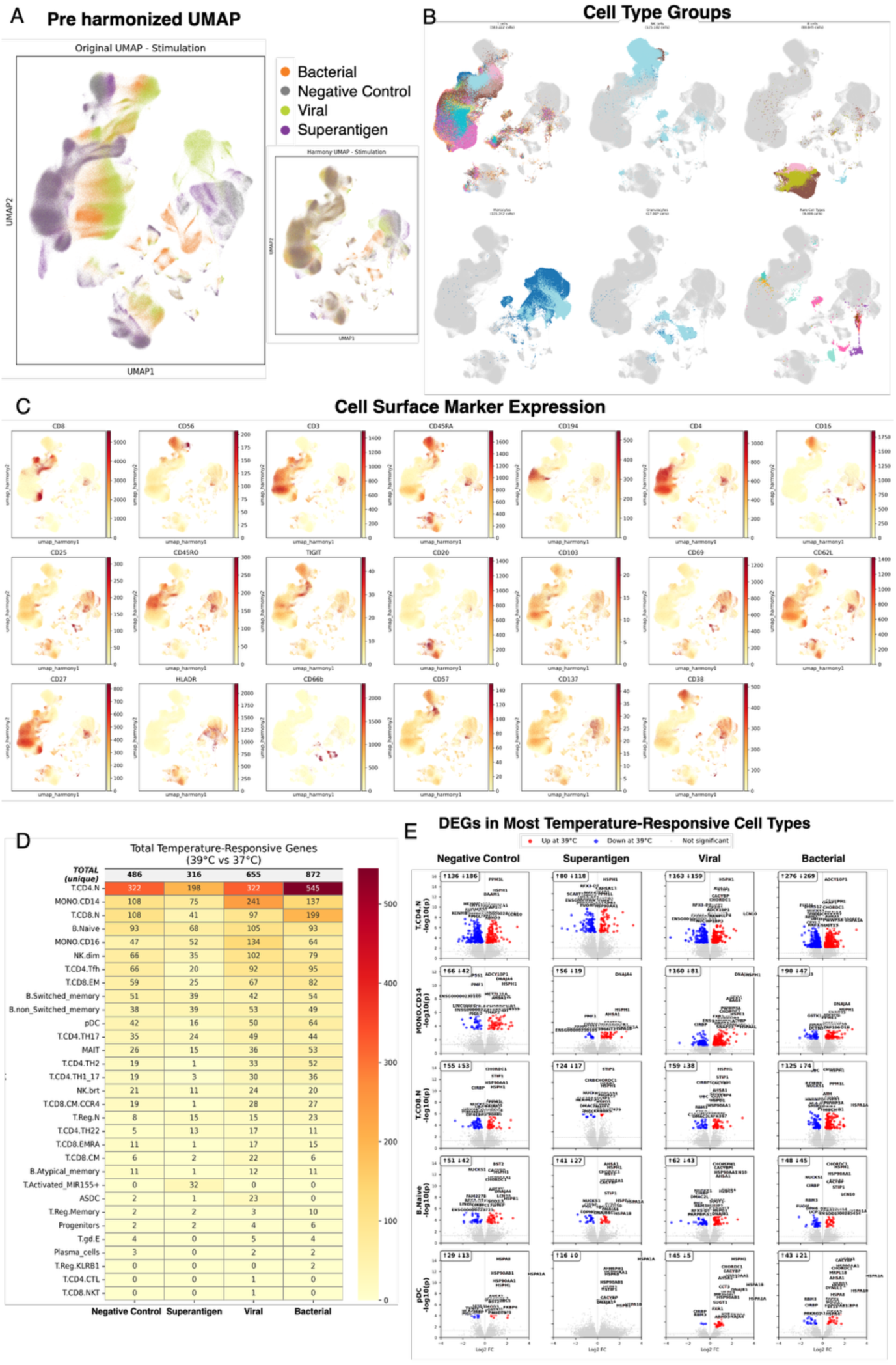
Single-cell analysis reveals cell type-specific temperature responses in *ex vivo* stimulated blood. **A** Pre-harmonized UMAP of 959,091 PBMCs colored by stimulation condition, (grey = negative control, purple = CytoStim, green = Poly I:C, orange = *E. coli*) with inset showing harmonized integration across stimulations **B** Cell type identification across 42 distinct populations spanning myeloid, B, T, and NK lineages, with individual UMAP plots highlighting each major cell group. **C** CITE-seq surface marker expression patterns across identified cell populations, showing antibody-derived protein levels for indicated immune cell markers. **D** Heatmap quantifying temperature-responsive genes (39°C vs 36.8-37°C) across cell types and stimulation conditions in sex aggregated analysis (n=6, 3 females, 3 males), with color intensity indicating the number of significantly differentially expressed genes per cell type-stimulus combination. **E** Volcano plots displaying differentially expressed genes (DEGs) in the most temperature-sensitive cell types across stimulation conditions, with significant genes (FDR corrected q-value < 0.05, |log2FC| > 0.25) highlighted and top 25 most significant labeled.

**Supplementary Figure 5.**
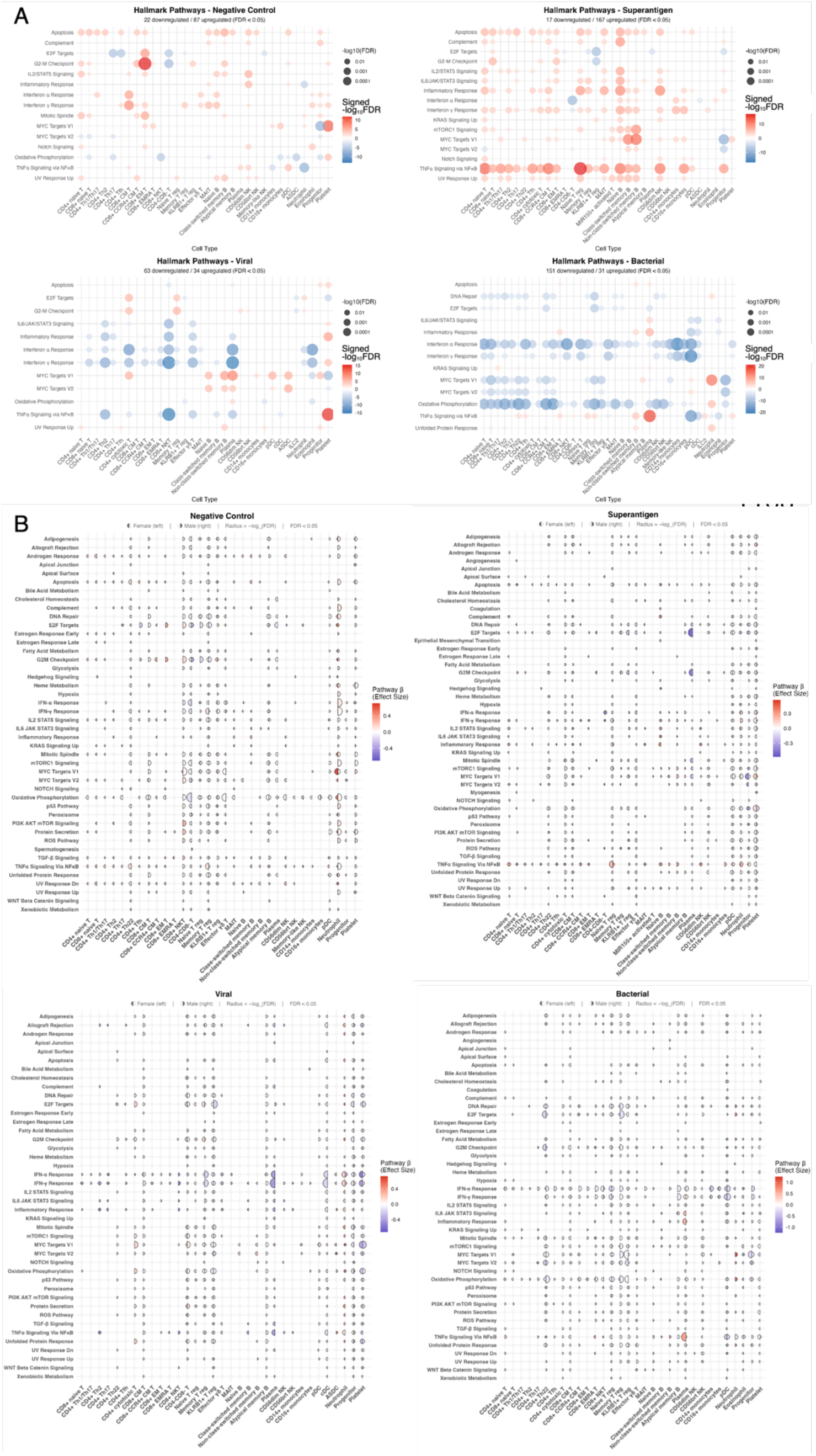
Single-cell pseudobulk temperature-enriched Hallmark gene sets. Hallmark gene set test analysis showing the temperature-responsive gene sets across cell types for the indicated stimulation condition in **A** all donors and **B** with the sexes analyzed independently. Dot size indicates −log10(FDR) and color represents direction of regulation (red = upregulated, blue = downregulated at 39°C vs 36.8°C). In B, female responses are the left hemisphere and male responses are the right hemisphere.

**Supplementary Figure 6.**
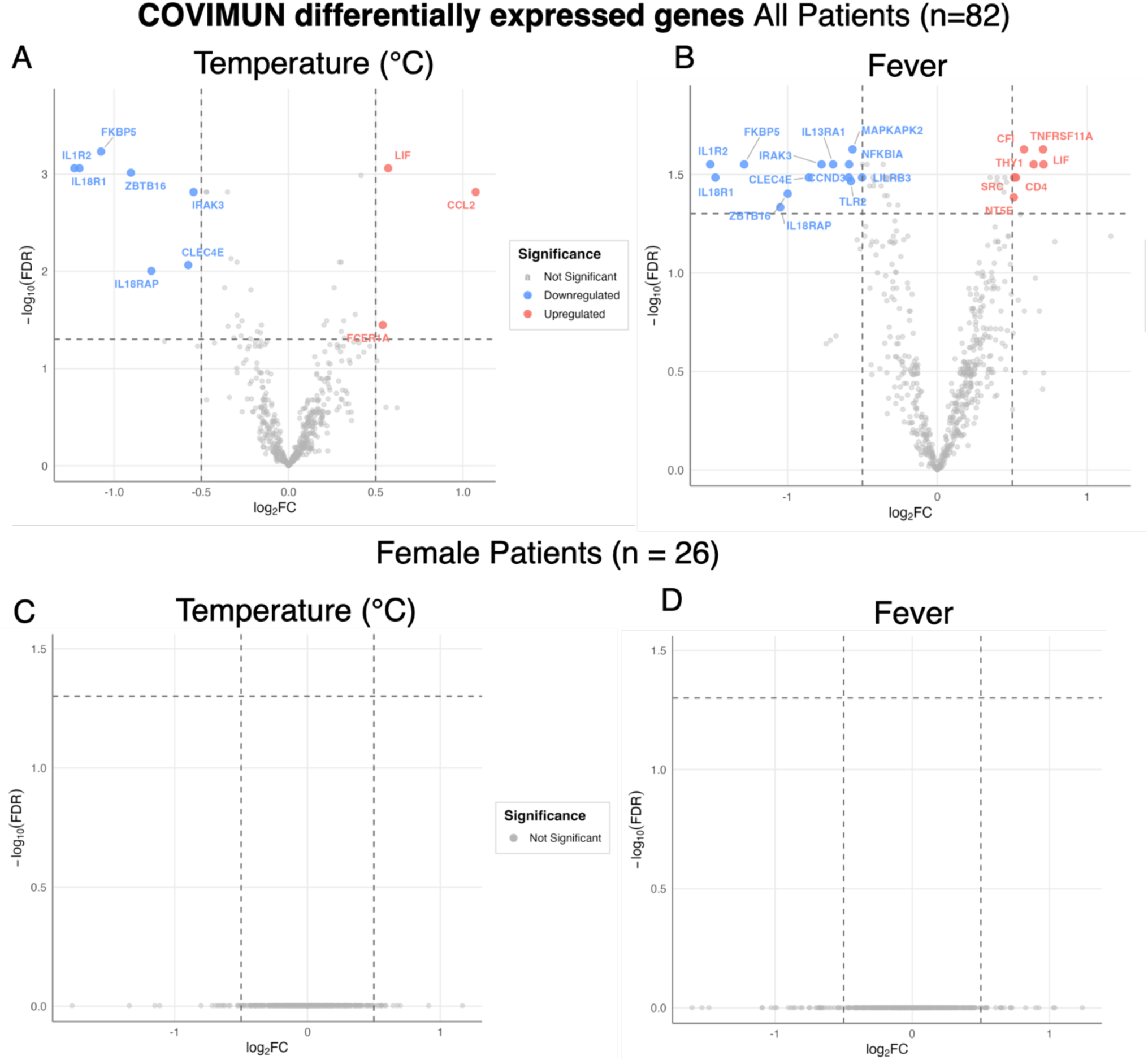
COVIMUN differentially expressed genes in all patients and female subcohort. Volcano plots of differentially expressed genes in all COVIMUN donors (n=82) associated with **A** temperature and **B** fever, and in female patients (n = 26) associated with **C** temperature and **D** fever.

**Supplementary Figure 7.**
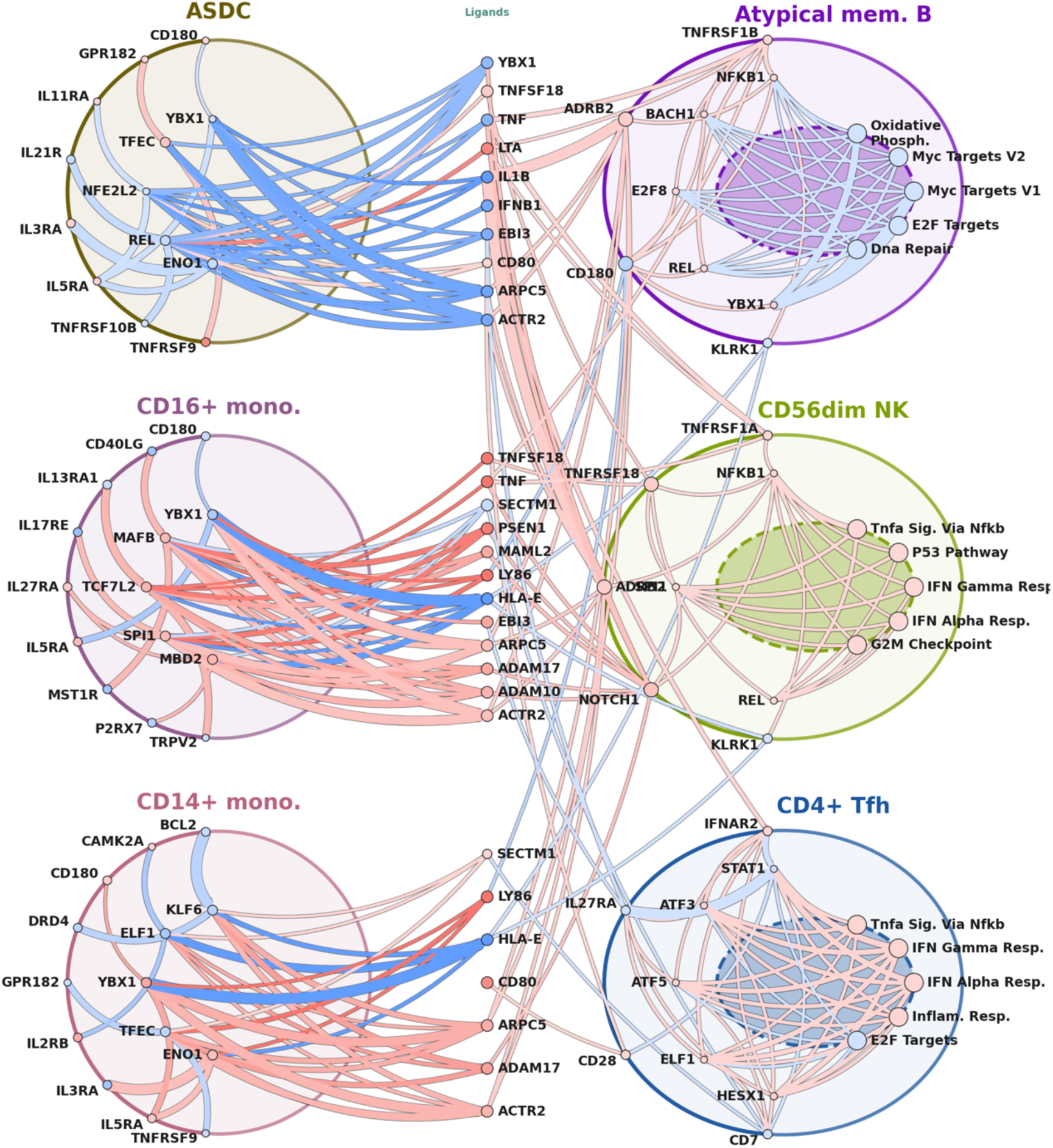
ReCoN viral stimulation cascade plot in female donors. Pseudotime cascade plots for the female viral stimulation condition showing temperature-modulated cell-to-cell communication. The left column shows the 3 “sender cells”, the middle column their differentially expressed molecular signals being sent to the receiver cells in the right column. Dots on the sender cell membranes represent the top receptors through which they are being regulated, signaling to the cytoplasm dots representing the top transcription factors modulating differentially expressed ligand genes shown in the column between the sender cells and receiver cell. These ligands are recognized by the receptor dots on the receiver cell membrane, signaling to the transcription factors represented by dots in the cytoplasm, and modulating gene expression in most significantly modulated Hallmark gene sets for that cell. Dot and connector color are determined by the temperature effect on gene expression or ReCoN score (red = 39°C, blue = 36.8°C).

**Supplementary Table 1.**
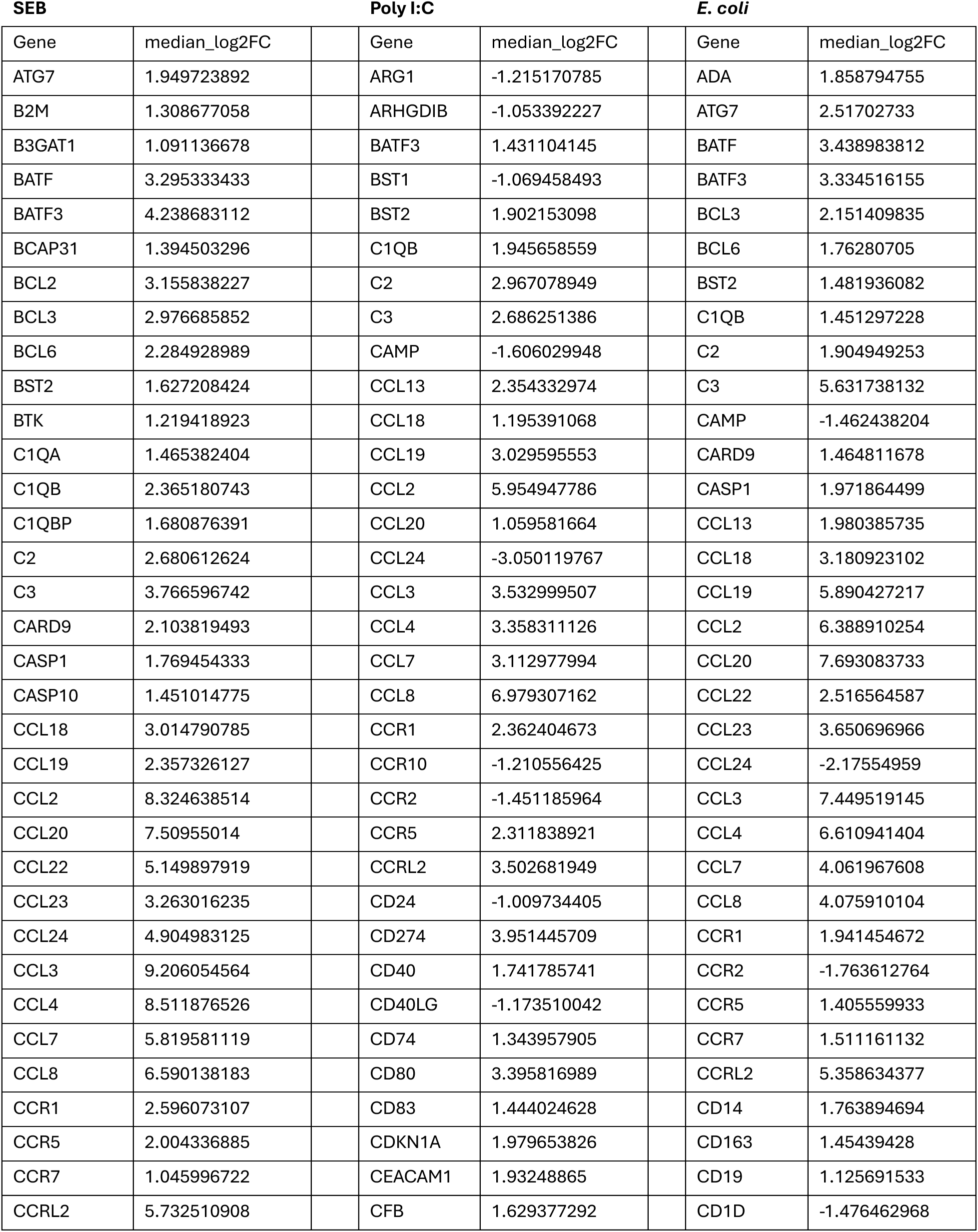
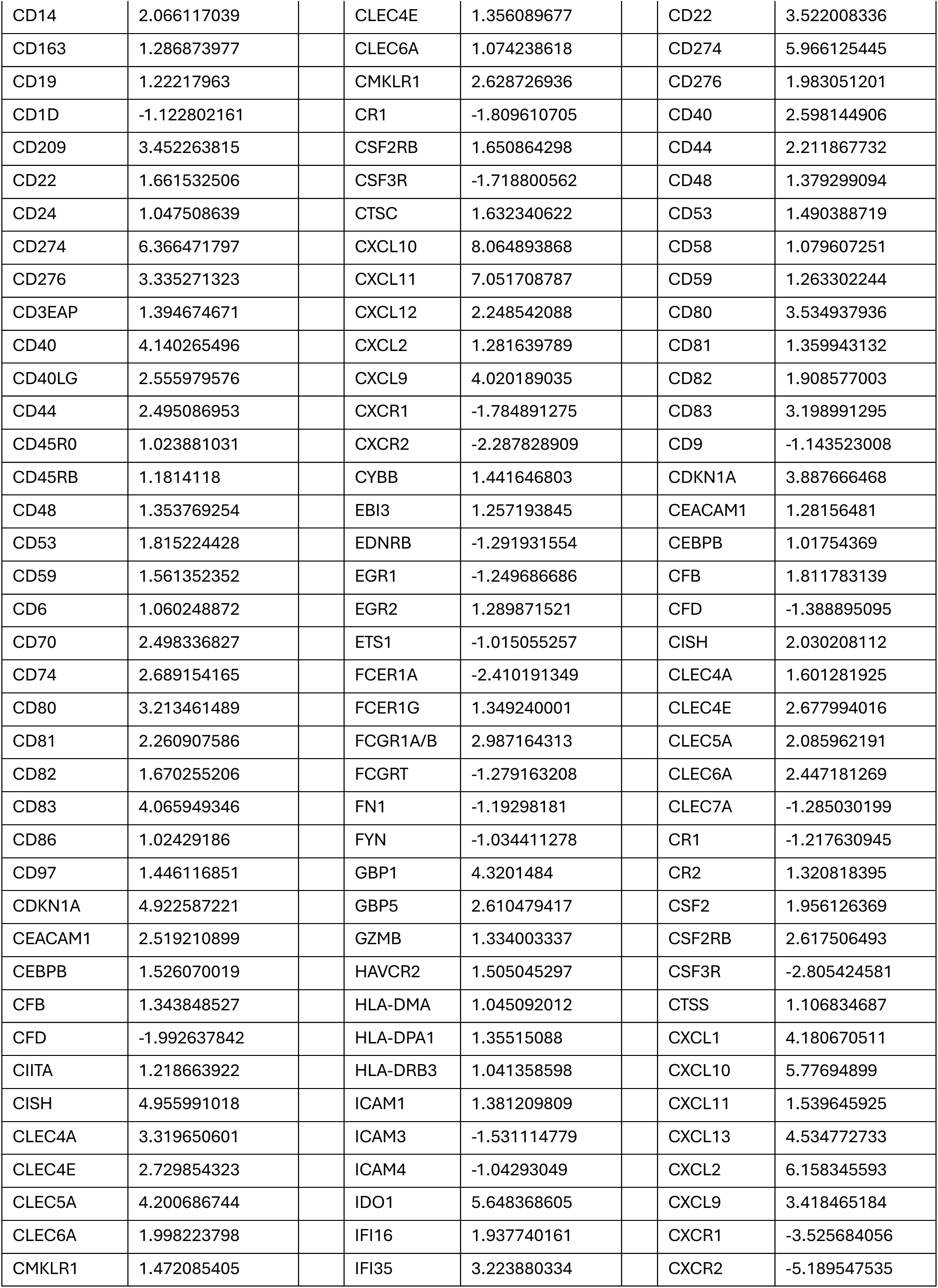
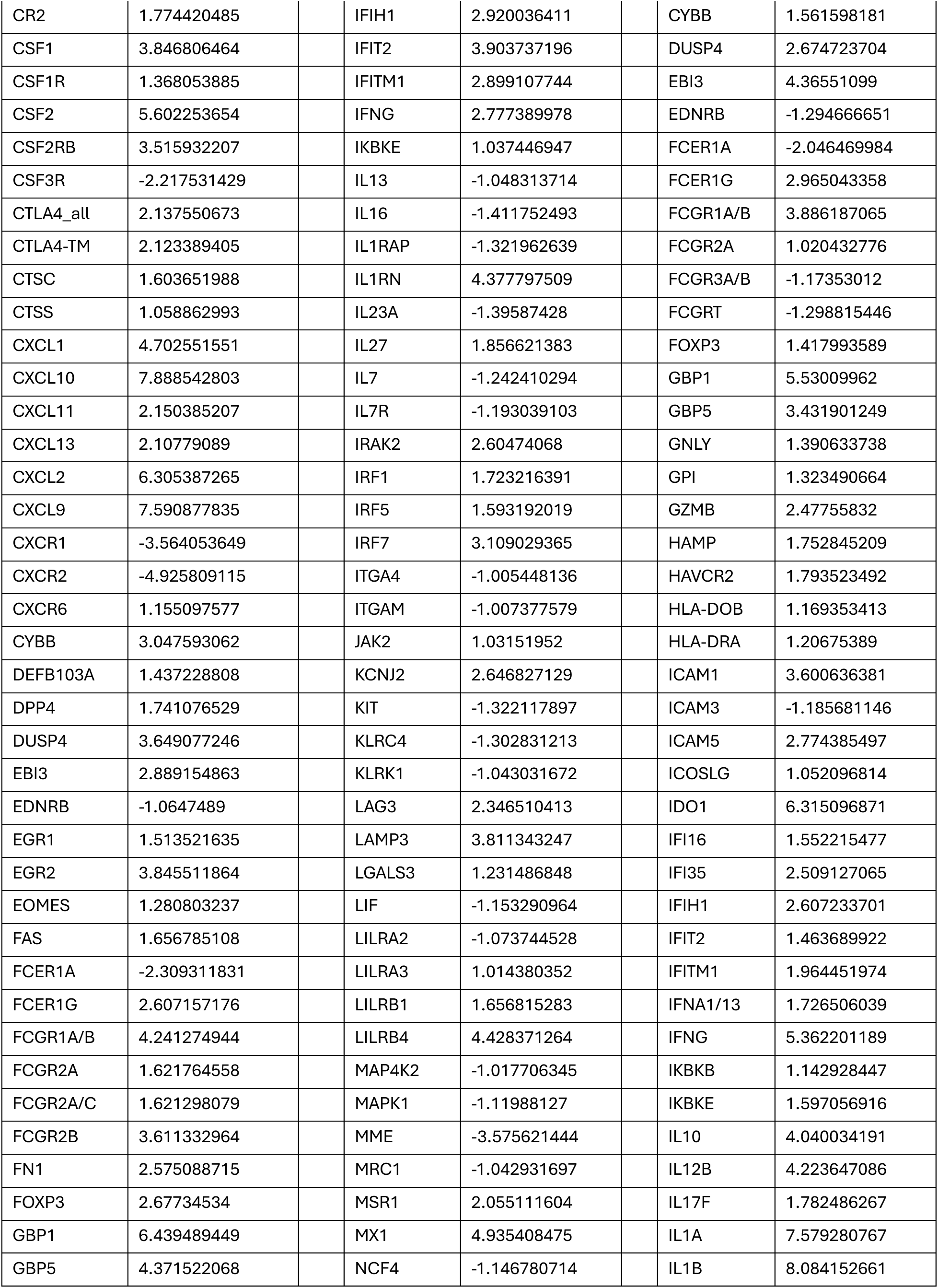
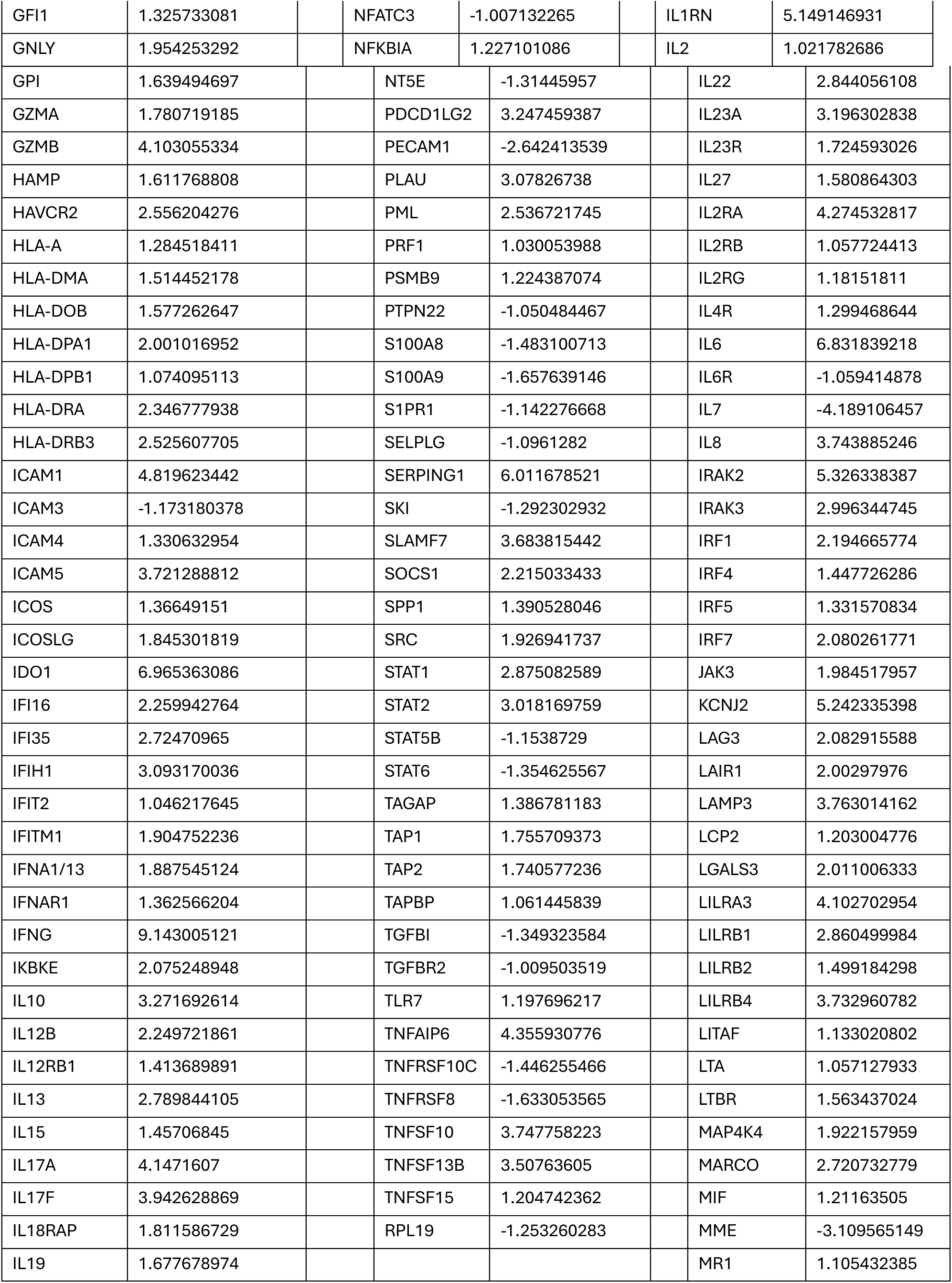
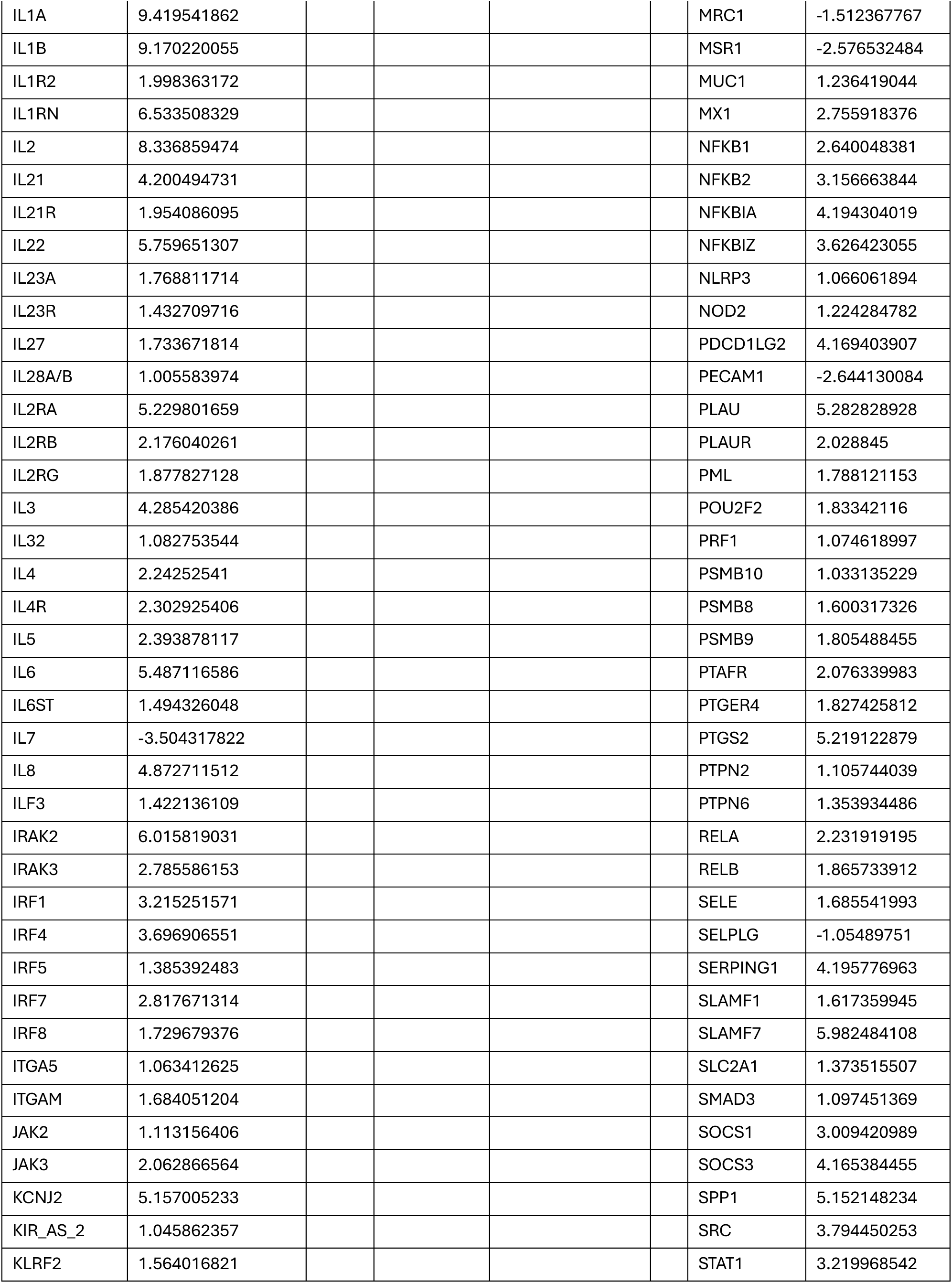
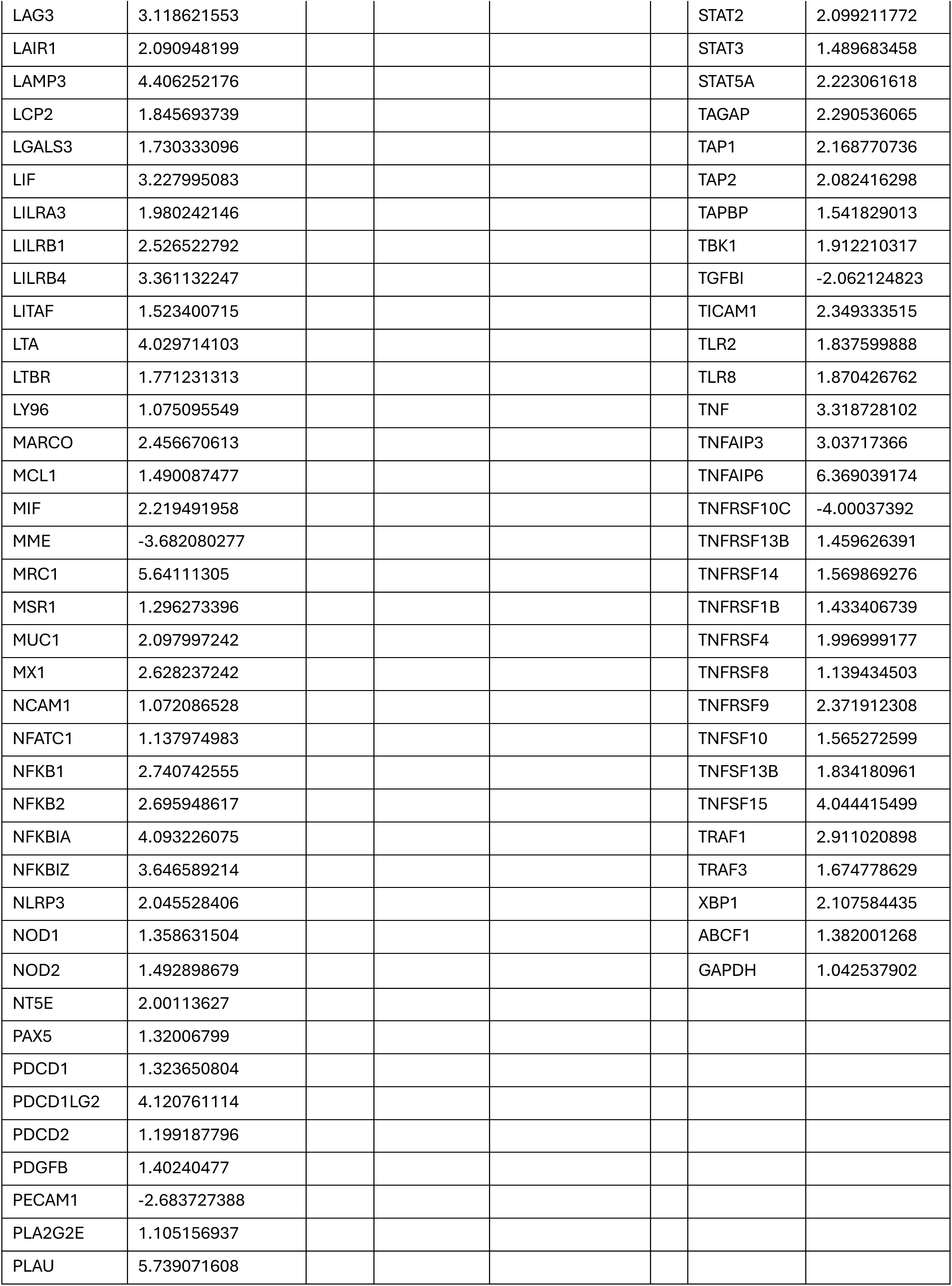
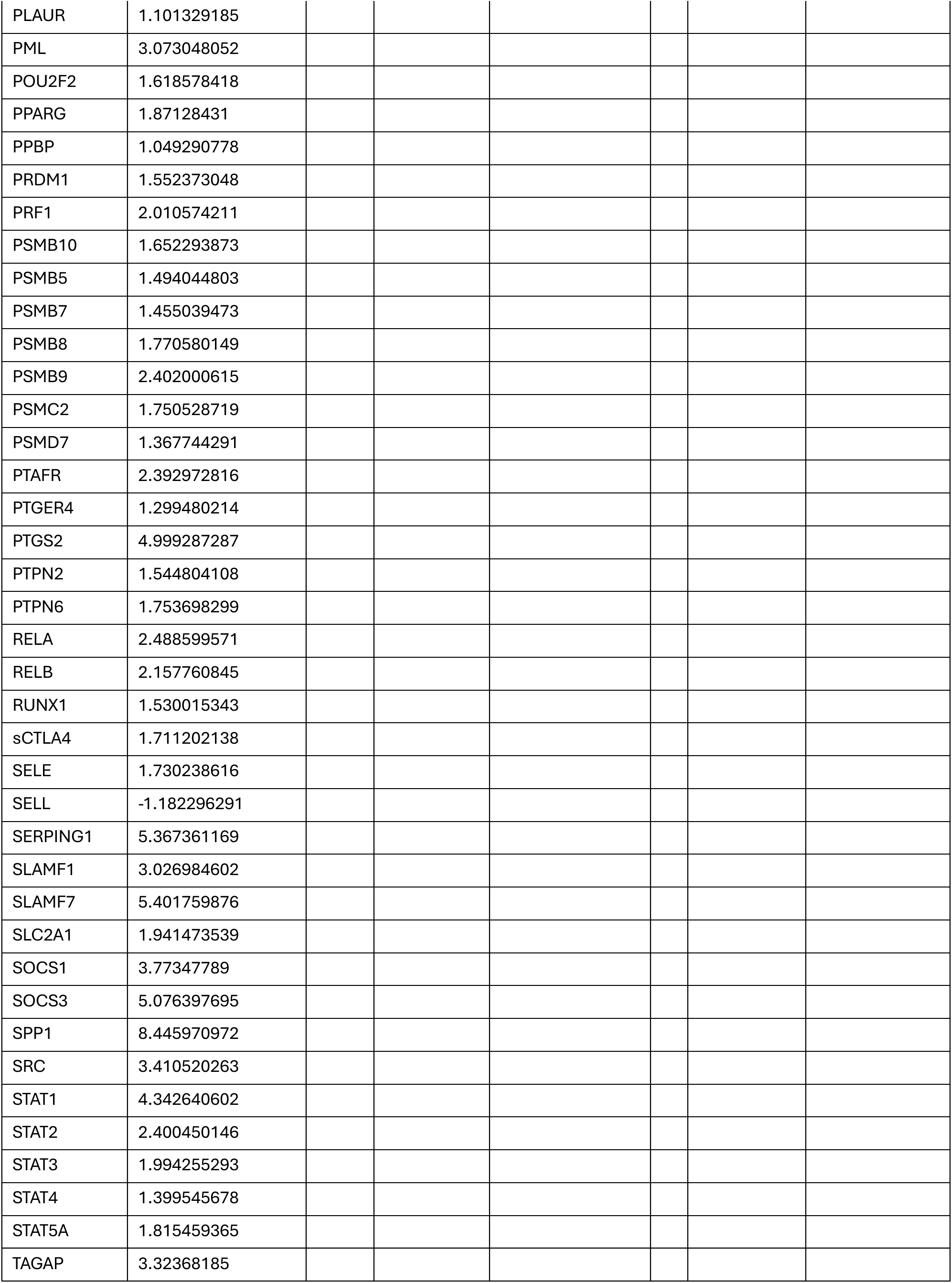
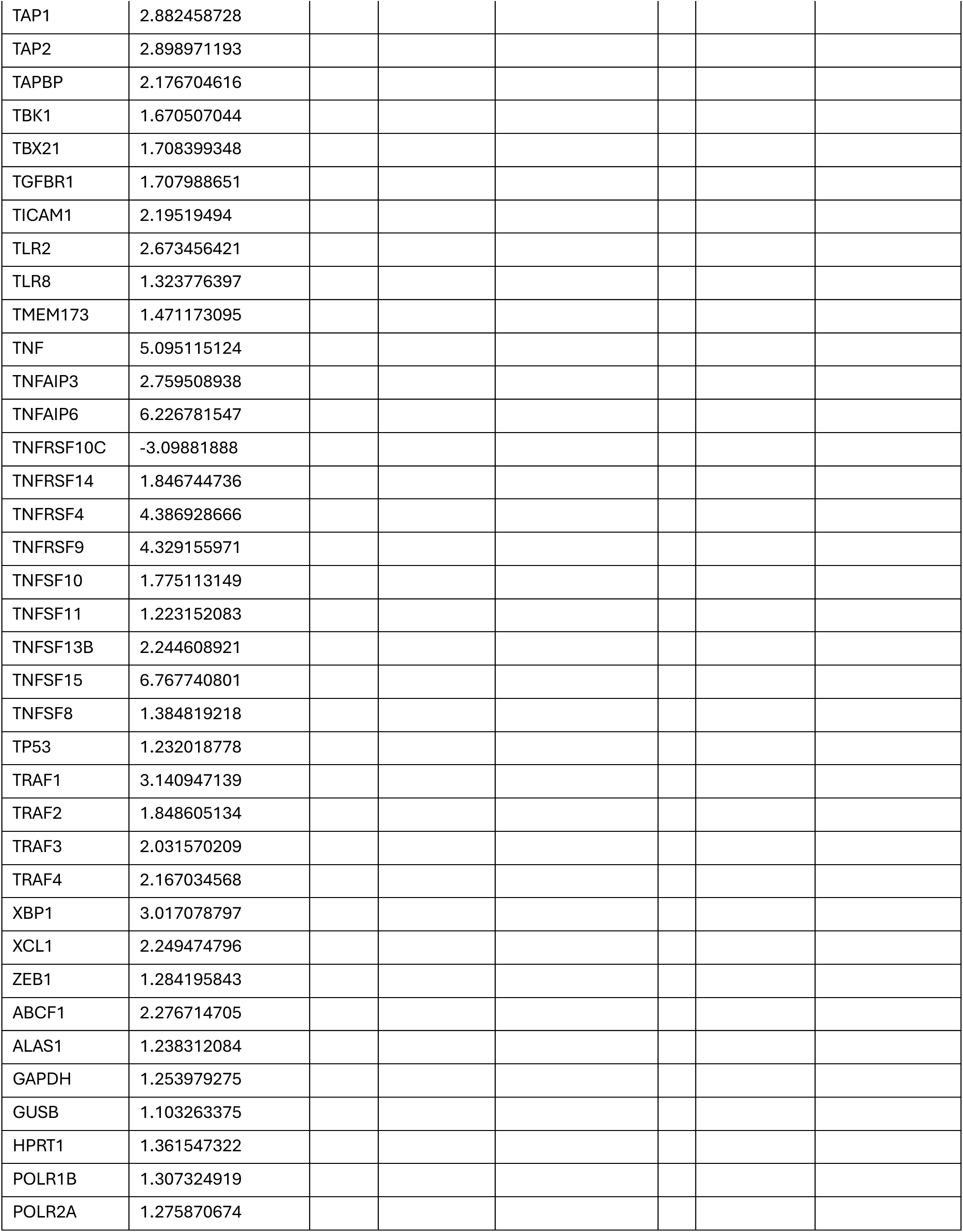
Results from MI *ex vivo* induced transcriptional response regression models. For each gene whose expression was measured by NanoString, the median log2FC of the indicated stimulated condition over the nonstimulated condition in the MI cohort is shown. These log2FC values were the response variables in the transcriptional response regression models in Figures 2, 3.

**Supplementary Table 2.** COVIMUN All Patients Metadata.

|  | **All**<br>N = 82 |  |
| --- | --- | --- |
| **Age, years median (IQR)** | 65 | (57 - 75) |
| **Age group, years n (%)** |  |  |
| **18 - 39** | 6 | -7.3 |
| **40 – 64** | 28 | -34 |
| **> 65** | 48 | -59 |
| **Female, n (%)** | 26 | -32 |
| **Body Mass Index, kg/m2, median (IQR)** | 29 | (24 - 32) |
| **Pregnancy, n (%)** | 1 | -1.2 |
| **Immunosuppressive condition or medicine, n (%)** | 22 | -27 |
| **Cancer, active, any, n (%)** | 16 | -20 |
| **Medicine, steroid before admission, n (%)** | 2 | -2.4 |
| **Surgical procedure, within 30 days pre-inclusion, n (%)** | 4 | -4.9 |
| **Vaccinated, COVID-19, 1st dose prior to admission, n (%)** | 2 | -2.4 |
| **Peak severity grade, n (%)** |  |  |
| **Grade I (< 3 L O2 for SpO2 > 92%)** | 30 | -37 |
| **Grade II (> 3 L O2 for SpO2 > 92%)** | 20 | -24 |
| **Grade III (> 6 L O2 for SpO2 > 92%)** | 19 | -23 |
| **Grade IV (mechanical ventilation and/or death)** | 13 | -16 |
| **ICU admission, n (%)** | 16 | -20 |
| **Mechanical ventilation, n (%)** | 11 | -13 |
| **Death, during admission, n (%)** | 6 | -7.3 |
| **Death, < 90-day mortality, n (%)** | 9 | -11 |

**Supplementary Table 3.** TotalSeq^TM^-B references for the antibodies used in the CITE-seq experiments.

| **Product Code** | **Product** | **Clone** | **Supplier** | **Dilution** |
| --- | --- | --- | --- | --- |
| 309837 | TotalSeq™-B0355 anti-human CD137 (4-1BB) | 4B4-1 | Biolegend | 1/50 |
| 307661 | TotalSeq™-B0159 anti-human HLA-DR | L243 | Biolegend | 1/50 |
| 302063 | TotalSeq™-B0083 anti-human CD16 | 3G8 | Biolegend | 1/50 |
| 302361 | TotalSeq™-B0100 anti-human CD20 | 2H7 | Biolegend | 1/50 |
| 362561 | TotalSeq™-B0047 anti-human CD56 (NCAM) | 5.1H11 | Biolegend | 1/50 |
| 302647 | TotalSeq™-B0085 anti-human CD25 | BC96 | Biolegend | 1/50 |
| 350235 | TotalSeq™-B0145 anti-human CD103 (Integrin αE) | Ber-ACT8 | Biolegend | 1/50 |
| 304849 | TotalSeq™-B0147 anti-human CD62L | DREG-56 | Biolegend | 1/50 |
| 310949 | TotalSeq™-B0146 anti-human CD69 | FN50 | Biolegend | 1/50 |
| 303547 | TotalSeq™-B0389 anti-human CD38 | HIT2 | Biolegend | 1/50 |
| 304161 | TotalSeq™-B0063 anti-human CD45RA | HI100 | Biolegend | 1/50 |
| 302851 | TotalSeq™-B0154 anti-human CD27 | O323 | Biolegend | 1/50 |
| 393323 | TotalSeq™-B0168 anti-human CD57 Recombinant Antibody | QA17A04 | Biolegend | 1/50 |
| 300565 | TotalSeq™-B0072 anti-human CD4 | RPA-T4 | Biolegend | 1/50 |
| 344757 | TotalSeq™-B0046 anti-human CD8 | SK1 | Biolegend | 1/50 |
| 344853 | TotalSeq™-B0049 anti-human CD3 | SK7 | Biolegend | 1/50 |
| 304257 | TotalSeq™-B0087 anti-human CD45RO | UCHL1 | Biolegend | 1/50 |
| 372727 | TotalSeq™-B0089 anti-human TIGIT (VSTM3) | A15153G | Biolegend | 1/50 |
| 359427 | TotalSeq™-B0071 anti-human CD194 (CCR4) | I.291H4 | Biolegend | 1/50 |
| 392913 | TotalSeq™-B0166 anti-human CD66b | 6/40c | Biolegend | 1/50 |

## List of Acronyms

ASDC: AXL^+^ SIGLEC6^+^ dendritic cell
CD: cluster of differentiation
CITE-seq: cellular indexing of transcriptome and epitope sequencing
CM: central memory
CMV: cytomegalovirus
CXCL: chemokine ligand
*E. coli*: Escherichia coli
FC: fold change
FDR: false discovery rate
G-CSF: granulocyte colony stimulating factor
GM-CSF: granulocyte-macrophage colony-stimulating factor
HSP: heat shock proteins
HSR: heat shock response
IFN: interferon
IL: interleukin
LPS: lipopolysaccharides
LRT: likelihood ratio testing
MAIT: mucosal-associated invariant T cells
MAPK: mitogen activated protein kinase
MDA-5: melanoma differentiation-associate gene 5
MI: *Milieu Intérieur*
NF-κB: nuclear factor kappa light chain enhancer of activated B cells
NK: natural killer cell
NLRP3: NOD like receptor protein 3
PBMC: peripheral blood mononuclear cell
pDC: plasmacytoid dendritic cell
Poly I:C: polyinosinic:polycytidylic acid
R848: resiquimod
RIG-I: retinoic acid inducible protein I
SEB: staphylococcal enterotoxin B
TLR: toll like receptor
TNF: tumor necrosis factor

## References

1. Mauvais-Jarvis, F. et al. Sex and gender: modifiers of health, disease, and medicine. The Lancet 396, 565–582 (2020).

2. Gay, L. et al. Sexual Dimorphism and Gender in Infectious Diseases. Front. Immunol. 12, (2021).

3. Klein, S. L. & Flanagan, K. L. Sex differences in immune responses. Nat. Rev. Immunol. 16, 626–638 (2016).

4. Piasecka, B., et al. Distinctive roles of age, sex, and genetics in shaping transcriptional variation of human immune responses to microbial challenges. Proc. Natl. Acad. Sci. 115, E488–E497 (2018).

5. Syrett, C. M. & Anguera, M. C. When the balance is broken: X-linked gene dosage from two X chromosomes and female-biased autoimmunity. J. Leukoc. Biol. 106, 919–932 (2019).

6. Migeon, B. R. X-linked diseases: susceptible females. Genet. Med. 22, 1156–1174 (2020).

7. Wira, C. R. & Fahey, J. V. A new strategy to understand how HIV infects women: identification of a window of vulnerability during the menstrual cycle. AIDS Lond. Engl. 22, 1909–1917 (2008).

8. Ikuta, K., Ejima, A., Abe, S. & Shimba, A. Control of immunity and allergy by steroid hormones. Allergol. Int. 71, 432–436 (2022).

9. Lakshmikanth, T. et al. Immune system adaptation during gender-affirming testosterone treatment. Nature 633, 155–164 (2024).

10. Maloney, E. & Duffy, D. Deciphering the relationship between temperature and immunity. Discov. Immunol. 3, kyae001 (2024).

11. Ley, C. et al. Defining Usual Oral Temperature Ranges in Outpatients Using an Unsupervised Learning Algorithm. JAMA Intern. Med. https://doi.org/10.1001/jamainternmed.2023.4291 (2023) doi:10.1001/jamainternmed.2023.4291.

12. Bolhassani, A. & Agi, E. Heat shock proteins in infection. Clin. Chim. Acta 498, 90–100 (2019).

13. Hasday, J. & Singh, I. Fever and the heat shock response: Distinct, partially overlapping processes. Cell Stress Chaperones 5, 471–80 (2000).

14. Murapa, P., Gandhapudi, S., Skaggs, H. S., Sarge, K. D. & Woodward, J. G. Physiological Fever Temperature Induces a Protective Stress Response in T Lymphocytes Mediated by Heat Shock Factor-1 (HSF1)1. J. Immunol. 179, 8305–8312 (2007).

15. van Eden, W. et al. The Enigma of Heat Shock Proteins in Immune Tolerance. Front. Immunol. 8, 1599 (2017).

16. Vabulas, R. M., Wagner, H. & Schild, H. Heat Shock Proteins as Ligands of Toll-Like Receptors. in Toll-Like Receptor Family Members and Their Ligands (eds. Beutler, B. & Wagner, H.) 169–184 (Springer, Berlin, Heidelberg, 2002). doi:10.1007/978-3-642-59430-4_11.

17. Hu, C. et al. Heat shock proteins: Biological functions, pathological roles, and therapeutic opportunities. MedComm 3, e161 (2022).

18. Barna, J., Csermely, P. & Vellai, T. Roles of heat shock factor 1 beyond the heat shock response. Cell. Mol. Life Sci. 75, 2897–2916 (2018).

19. van Eden, W., Spiering, R., Broere, F. & van der Zee, R. A case of mistaken identity: HSPs are no DAMPs but DAMPERs. Cell Stress Chaperones 17, 281–292 (2012).

20. Tarner, I. H., Müller-Ladner, U., Uhlemann, C. & Lange, U. The effect of mild whole-body hyperthermia on systemic levels of TNF-alpha, IL-1beta, and IL-6 in patients with ankylosing spondylitis. Clin. Rheumatol. 28, 397–402 (2009).

21. Kida, Y. et al. Increased liver temperature efficiently augments human cellular immune response: T-cell activation and possible monocyte translocation. Cancer Immunol. Immunother. CII 55, 1459–1469 (2006).

22. Jeziorski, K. Hyperthermia in rheumatic diseases. A promising approach? Reumatologia 56, 316–320 (2018).

23. Basu, S. & Srivastava, P. K. Fever-like temperature induces maturation of dendritic cells through induction of hsp90. Int. Immunol. 15, 1053–1061 (2003).

24. Wang, X. et al. Febrile Temperature Critically Controls the Differentiation and Pathogenicity of T Helper 17 Cells. Immunity 52, 328–341.e5 (2020).

25. Evans, S. S., Repasky, E. A. & Fisher, D. T. Fever and the thermal regulation of immunity: the immune system feels the heat. Nat. Rev. Immunol. 15, 335–349 (2015).

26. Saint-André, V. et al. Smoking changes adaptive immunity with persistent effects. Nature 626, 827–835 (2024).

27. Deltourbe, L. G. et al. Steroid hormone levels vary with sex, aging, lifestyle, and genetics. 2024.10.07.24315000 Preprint at 10.1101/2024.10.07.24315000 (2024).

28. Heal, C., Harvey, A., Brown, S., Rowland, A. G. & Roland, D. The association between temperature, heart rate, and respiratory rate in children aged under 16 years attending urgent and emergency care settings. Eur. J. Emerg. Med. 29, 413–416 (2022).

29. Kirschen, G. W., Singer, D. D., Thode, H. C. & Singer, A. J. Relationship between body temperature and heart rate in adults and children: A local and national study. Am. J. Emerg. Med. 38, 929–933 (2020).

30. Patin, E. et al. Natural variation in the parameters of innate immune cells is preferentially driven by genetic factors. Nat. Immunol. 19, 302–314 (2018).

31. Duffy, D. et al. Functional Analysis via Standardized Whole-Blood Stimulation Systems Defines the Boundaries of a Healthy Immune Response to Complex Stimuli. Immunity 40, 436–450 (2014).

32. Urrutia, A. et al. Standardized Whole-Blood Transcriptional Profiling Enables the Deconvolution of Complex Induced Immune Responses. Cell Rep. 16, 2777–2791 (2016).

33. Saint-André, V. et al. Smoking changes adaptive immunity with persistent effects. Nature 626, 827–835 (2024).

34. Stoeckius, M. et al. Simultaneous epitope and transcriptome measurement in single cells. Nat. Methods 14, 865–868 (2017).

35. Soto-Heredero, G. et al. KLRG1 identifies regulatory T cells with mitochondrial alterations that accumulate with aging. Nat. Aging 5, 799–815 (2025).

36. Zheng, M. et al. TLR2 senses the SARS-CoV-2 envelope protein to produce inflammatory cytokines. Nat. Immunol. 22, 829–838 (2021).

37. Khabar, K. S. A. Post-transcriptional control during chronic inflammation and cancer: a focus on AU-rich elements. Cell. Mol. Life Sci. 67, 2937–2955 (2010).

38. Trimbour, R., Flores, R. O. R., Saez-Rodriguez, J. & Cantini, L. Modelling multicellular coordination by bridging cell-cell communication and intracellular regulation through multilayer networks. 2026.01.20.700561 Preprint at 10.64898/2026.01.20.700561 (2026).

39. Inouye, S. et al. Heat shock-induced heme oxygenase-1 expression in a mouse hepatoma cell line is dependent on HSF1 and modified by NRF2 and BACH1. Genes Cells 27, 719–730 (2022).

40. Ahmadivand, S., Fux, R. & Palić, D. Role of T Follicular Helper Cells in Viral Infections and Vaccine Design. Cells 14, 508 (2025).

41. Priest, D. G. et al. Atypical and non-classical CD45RBlo memory B cells are the majority of circulating SARS-CoV-2 specific B cells following mRNA vaccination or COVID-19. Nat. Commun. 15, 6811 (2024).

42. Oliviero, B. et al. Expansion of atypical memory B cells is a prominent feature of COVID-19. Cell. Mol. Immunol. 17, 1101–1103 (2020).

43. Portugal, S., Obeng-Adjei, N., Moir, S., Crompton, P. D. & Pierce, S. K. Atypical memory B cells in human chronic infectious diseases: An interim report. Cell. Immunol. 321, 18–25 (2017).

44. Obermeyer, Z., Samra, J. K. & Mullainathan, S. Individual differences in normal body temperature: longitudinal big data analysis of patient records. BMJ 359, j5468 (2017).

45. Hsieh, C.-S. & Bautista, J. L. Sliding set-points of immune responses for therapy of autoimmunity. J. Exp. Med. 207, 1819–1823 (2010).

46. Lei, Y. & Tsang, J. S. Systems Human Immunology and AI: Immune Setpoint and Immune Health.

47. Wang, Wei et al. NLRP3 is a thermosensor that is negatively regulated by high temperature. in vol. 54 143 (Eur. J. Immunol., 2024).

48. Manuel, R. S. J. & Liang, Y. Sexual Dimorphism in Immunometabolism and Autoimmunity: Impact on Personalized Medicine. Autoimmun. Rev. 20, 102775 (2021).

49. Metcalf, C. J. E. & Graham, A. L. Schedule and magnitude of reproductive investment under immune trade-offs explains sex differences in immunity. Nat. Commun. 9, 4391 (2018).

50. Bjorklund, D. F. & Shackelford, T. K. Differences in Parental Investment Contribute to Improvement Differences between Men and Women. Curr. Dir. Psychol. Sci. 8, 86–89 (1999).

51. Mitchell, E., Graham, A. L., Úbeda, F. & Wild, G. On maternity and the stronger immune response in women. Nat. Commun. 13, 4858 (2022).

52. Lakshmikanth, T. et al. Immune system adaptation during gender-affirming testosterone treatment. Nature 633, 155–164 (2024).

53. Aibiki, M., Maekawa, S., Nishiyama, T., Seki, K. & Yokono, S. Activated cytokine production in patients with accidental hypothermia. Resuscitation 41, 263–268 (1999).

54. Welzel, T. & Kuemmerle-Deschner, J. B. Diagnosis and Management of the Cryopyrin-Associated Periodic Syndromes (CAPS): What Do We Know Today? J. Clin. Med. 10, 128 (2021).

55. Henderson, M. E. T. & Halsey, L. G. The metabolic upper critical temperature of the human thermoneutral zone. J. Therm. Biol. 110, 103380 (2022).

56. Ocha, I. Extreme Heat: Preparing for the Heatwaves of the Future. Oct. 2022.

57. Spiljar, M. et al. Cold exposure protects from neuroinflammation through immunologic reprogramming. Cell Metab. 33, 2231–2246.e8 (2021).

58. Koch Esteves, N., Gibson, O. R., Khir, A. W. & González-Alonso, J. Regional thermal hyperemia in the human leg: Evidence of the importance of thermosensitive mechanisms in the control of the peripheral circulation. Physiol. Rep. 9, e14953 (2021).

59. Gupta, A. K., Wang, T., Cooper, E. A., Conenello, R. M. & Bristow, I. R. The treatment of plantar warts using microwave—A review of 85 consecutive cases in the United States. J. Cosmet. Dermatol. 22, 2729–2736 (2023).

60. Thomas, S. et al. The Milieu Intérieur study — An integrative approach for study of human immunological variance. Clin. Immunol. 157, 277–293 (2015).

61. Svanberg, R. et al. Early stimulated immune responses predict clinical disease severity in hospitalized COVID-19 patients. Commun. Med. 2, 114 (2022).

62. Bhattacharya, A. et al. An approach for normalization and quality control for NanoString RNA expression data. Brief. Bioinform. 22, bbaa163 (2020).

63. Risso, D., Ngai, J., Speed, T. P. & Dudoit, S. Normalization of RNA-seq data using factor analysis of control genes or samples. Nat. Biotechnol. 32, 896–902 (2014).

64. Law, C. W. et al. RNA-seq analysis is easy as 1-2-3 with limma, Glimma and edgeR. F1000Research 5, ISCB Comm J-1408 (2018).

65. GSEA | MSigDB | Human MSigDB Collections. https://www.gsea-msigdb.org/gsea/msigdb/human/collections.jsp#H.

66. Xu, C. et al. Automatic cell-type harmonization and integration across Human Cell Atlas datasets. Cell 186, 5876–5891.e20 (2023).

67. Trimbour, R., Flores, R. O. R., Saez-Rodriguez, J. & Cantini, L. Modelling multicellular coordination by bridging cell-cell communication and intracellular regulation through multilayer networks. 2026.01.20.700561 Preprint at 10.64898/2026.01.20.700561 (2026).

68. Moerman, T. et al. GRNBoost2 and Arboreto: efficient and scalable inference of gene regulatory networks. Bioinformatics 35, 2159–2161 (2019).

69. Dimitrov, D. et al. LIANA+ provides an all-in-one framework for cell–cell communication inference. Nat. Cell Biol. 26, 1613–1622 (2024).

70. Efremova, M., Vento-Tormo, M., Teichmann, S. A. & Vento-Tormo, R. CellPhoneDB: inferring cell–cell communication from combined expression of multi-subunit ligand–receptor complexes. Nat. Protoc. 15, 1484–1506 (2020).

71. Gu, Z., Gu, L., Eils, R., Schlesner, M. & Brors, B. circlize implements and enhances circular visualization in R. Bioinformatics 30, 2811–2812 (2014).

72. Subramanian, A. et al. Gene set enrichment analysis: A knowledge-based approach for interpreting genome-wide expression profiles. Proc. Natl. Acad. Sci. 102, 15545–15550 (2005).

73. Wu, D. & Smyth, G. K. Camera: a competitive gene set test accounting for inter-gene correlation. Nucleic Acids Res. 40, e133 (2012).

